# Systemic and Mucosal Antibody Correlates of Protection Against Bordetella pertussis in a Controlled Human Infection Model

**DOI:** 10.64898/2026.06.18.26355881

**Authors:** Mehak Zahoor Khan, Yogita Kapoor, Evgenii Kliuchnikov, Thendral Selvam, Agness E. Lakudzala, Laura Fontana, MaryBeth Culp, Susan Hariri, May Elsherif, Doug Lauffenburger, Scott Halperin, Marcela F. Pasetti, Galit Alter, Boris Julg

**Author notes:** Co-corresponding authors Contact: Boris Julg, MD PhD, Ragon Institute of Mass General, MIT and Harvard, 600 Main Street, Cambridge, MA 02139, USA. These authors contributed equally (Mehak Zahoor Khan, Yogita Kapoor, Evgenii Kliuchnikov). Disclaimer: The findings and conclusions in this report are those of the authors and do not necessarily represent the official position of the Centers for Disease Control and Prevention, US Department of Health and Human Services.

## Abstract

**Background:** Despite high vaccination coverage, pertussis has resurged globally. Whole-cell (wP) and acellular (aP) pertussis vaccines induce distinct immune profiles, yet immune correlates of protection against infection and symptomatic disease remain incompletely defined. We leveraged a controlled human infection model (CHIM) to identify systemic and mucosal humoral signatures associated with resistance to *Bordetella pertussis*.

**Methods:** Adults with documented history of vaccination had previously been enrolled in a CHIM study and challenged intranasally with *B. pertussis* D420. For the present work, longitudinal serum and nasal wash samples were analyzed using systems serology to comprehensively profile antibody features. Multivariate modeling and network analyses were performed to define discriminatory immune features.

**Findings:** Baseline aP vaccine antigen–specific antibodies did not distinguish infection outcomes. In wP-primed individuals, protection from *B. pertussis* infection was associated with broad, high-magnitude, polyfunctional antibody responses targeting non–canonical antigens, including BrkA, TcfA, OmpP, OmlA, FauA, and Pal. Protective signatures associated with resistance to symptomatic disease in both vaccine groups were characterized by enhanced Fc-receptor-engaging antibody profiles with distinct antigenic patterns shaped by vaccine history. Importantly, while conventional aP vaccine antigens failed to reliably distinguish individuals susceptible to infection or symptom development, correlates generated by integrated serum and mucosal models based on select non-canonical antigens achieved near-perfect discrimination of infection and symptom outcomes, outperforming models restricted to aP-vaccine. antigens only.

**Interpretation:** Resistance to infection was largely restricted to wP-primed individuals and was associated with integrated systemic and mucosal antibody responses directed against antigens beyond those included in acellular vaccines. Protection from symptomatic disease in both vaccine groups was linked to distinct antibody response signatures, shaped by prior vaccination history. These findings indicate that immune mechanisms preventing infection differ from those limiting clinical disease and provide a framework for redesign of next-generation pertussis vaccines aimed at blocking infection and symptomatic disease.

**Funding:** This work was supported by Ragon Institute Sundry to G.A. and B.J, U19AI167899 to D.L., and US CDC through 75D30120R67837 to Dalhousie University and 75D30122C15467 to the University of Maryland, Baltimore.

## INTRODUCTION

*Bordetella pertussis* (Bp), the causative agent of whooping cough, is a highly contagious gram-negative respiratory pathogen transmitted via aerosols. Introduction of the whole-cell pertussis (wP) vaccine in the late 1940s reduced pertussis incidence and mortality by >90% in countries with high vaccine coverage. However, despite sustained vaccination coverage, pertussis has resurged globally over the past two decades^1^. Improved diagnostics and natural disease cyclicity contribute to this trend, but a key potential factor is the introduction of the acellular pertussis (aP) vaccines in the 1990s. aP vaccines contain purified Bp antigens - pertussis toxin toxoid, filamentous hemagglutinin, pertactin, and fimbrial proteins, adsorbed to alum^1^. “While aP vaccines are less reactogenic than wP vaccines and prevent severe disease, they induce short-lived immunity and fail to reliably prevent infection or transmission.^2,3^

The discordant protection afforded by wP and aP vaccines on pertussis control is rooted in the distinct immune response profiles they induce. Natural infection and wP vaccination induce robust Th1/Th17-biased cellular immunity alongside broad antibody responses^4–6^, whereas aP vaccines preferentially generate Th2-skewed responses.^3^ Although anti-pertussis toxin antibodies protect against severe disease, effective bacterial clearance appears to require Th1/Th17-driven mechanisms, including opsonizing antibodies, IgA, and activation of phagocytes at mucosal sites alongside the development of memory B cells and tissue-resident memory T cells in the respiratory tract^7–11^. However, the precise antigen specificities and immunologic mechanisms that collectively shape long-lived immunity remain incompletely defined. Understanding the immunological differences between aP and wP vaccines in terms of protection against infection could inform the redesign of aP vaccines to achieve robust immunity conferred by wP, while avoiding the safety concerns historically associated with wP formulations. Towards this, immunoproteomic analyses of convalescent serum and nasopharyngeal wash of infected baboons identified *B. pertussis* antigens beyond those included in acellular vaccines, including surface-exposed proteins essential for persistence in the airway and potentially relevant for preventing infection and transmission^12^. Understanding whether these antigens are targeted in the setting of wP in humans, or can collaborate with aP antigens via specific immune effector mechanisms remain incompletely understood, but could provide critical insights for immune correlates of protection against Bp.

To define the immune correlates of protection associated with resistance to Bp infection and symptom development, we performed comprehensive humoral immune profiling in a controlled human infection model (CHIM) of pertussis in adults previously vaccinated with wP or aP vaccines. Participants were challenged with increasing dosages of Bp D420 isolate, and serum and nasal samples were collected longitudinally (Fig. 1a). Antibody isotypes, subclasses, Fc-receptor binding profiles, antibody-dependent complement deposition, and antibody-dependent opsonophagocytic activity were profiled against a panel of antigens included in the aP vaccine, immunoproteomically-defined antigens^12^, and control antigens. Overall, this study identifies distinct humoral signatures associated with infection resistance in wP-primed individuals and symptom modulation in both vaccine recipients across serum and nasal mucosal surface. By combining a CHIM model with high-dimensional systems serology^13^, this work provides one of the most comprehensive dissections of humoral immunity to pertussis to date and establishes a rational foundation for next-generation vaccine aimed at achieving durable, transmission-blocking immunity.

**Figure 1.**
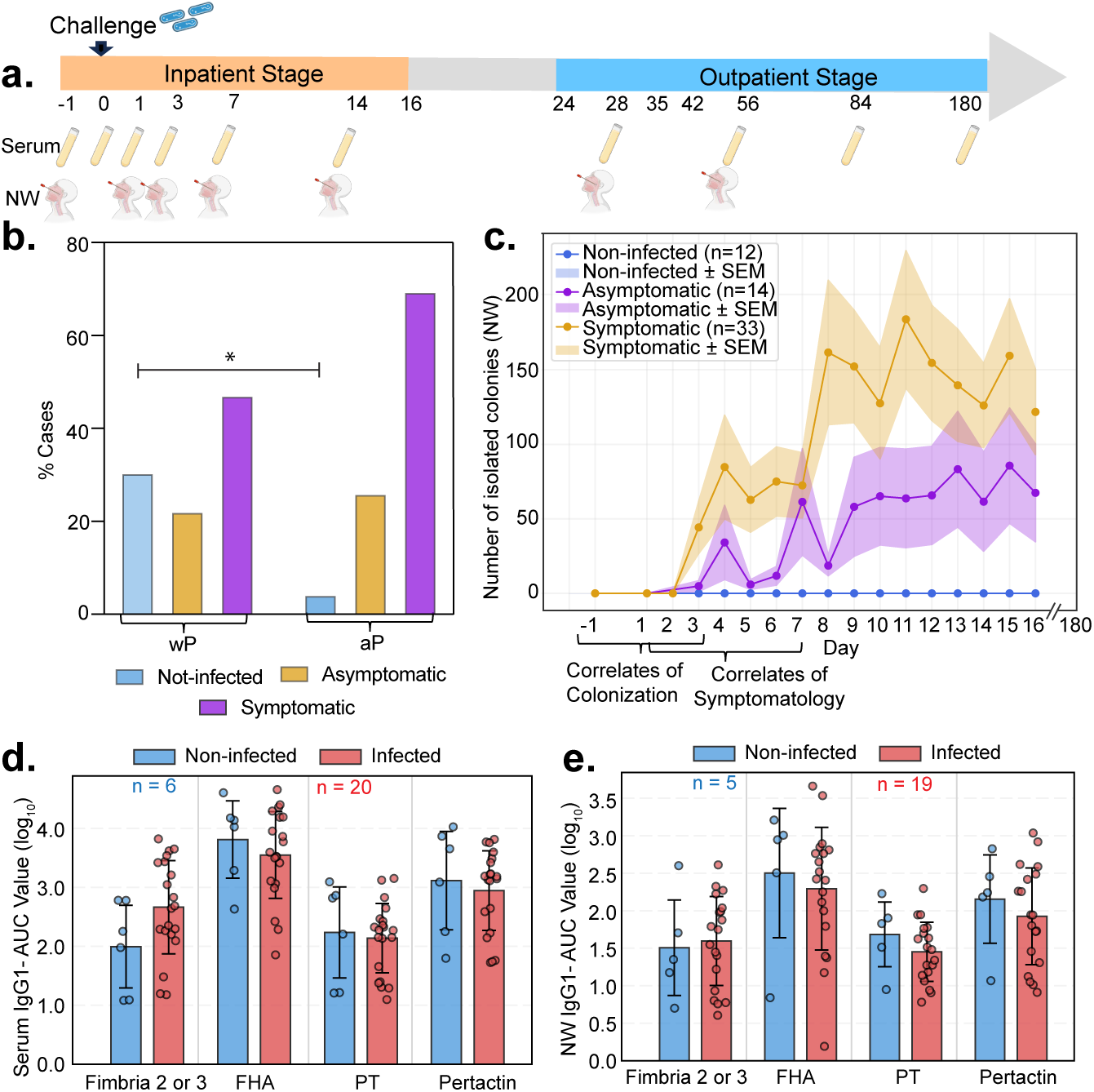
Study design, clinical outcomes and aP- antigen specific humoral responses in a controlled human infection model (CHIM) of Bordetella pertussis. (a) Study design and sampling timeline. Adult participants previously vaccinated with either whole-cell (wP) or acellular (aP) pertussis vaccines were challenged with B. pertussis D420 isolate and followed through inpatient and outpatient stages. Serum and nasal wash (NW) samples were collected longitudinally from baseline through day 180 post-challenge. (b) Distribution of clinical outcomes (See Methods) by vaccine history. Participants were categorized as non-infected, asymptomatic, or symptomatic following challenge, stratified by prior wP or aP vaccination. Significances were assessed using Fischer’s exact test. (c) The number of B. pertussis colonies isolated from nasal washes is shown for non-infected, asymptomatic, and symptomatic individuals (mean ± SEM). (d) Serum IgG1 responses to aP antigens, quantified as area under the curve (AUC) from day −1 to 3, comparing non-infected versus infected individuals. (e) NW IgG1 responses to aP antigens, quantified as area under the curve (AUC) from day −1 to 3, comparing non-infected versus infected individuals. The number of samples (n) from non-infected (blue) and infected (red) individuals is indicated in panels (e, f).

## RESULTS

### Clinical protection in the pertussis CHIM is not explained by baseline aP antigen–specific humoral immunity

To identify the correlates of protection against Bp infection, we leveraged data from the representing the first North American pertussis CHIM cohort (NCT05136599)^14^. In this model, healthy subjects aged 18-40 years with a documented history of vaccination with either wP or aP vaccines were challenged intranasally with a single dose inoculum of the Bp D420 clinical isolate and monitored longitudinally for infection, bacterial shedding, and clinical symptoms over a 180-day period. Participants were enrolled in a dose-escalation trial and received azithromycin treatment irrespective of clinical outcome (Fig. 1a), which was determined using a pre-specified automated algorithm incorporating quantitating bacterial infection and clinical symptomatology^14^.

Since CHIM provides a controlled framework to interrogate immune responses associated with Bp infection and disease in adults, we selected 451 serum samples and 318 nasal wash samples across 4 cohorts with challenge doses ranging from 10^6^ to 10^8^ CFU^14^, spanning multiple timepoints, enabling assessment of both baseline immunity and post-exposure immune dynamics. Clinical outcome assessment revealed marked differences between vaccine priming groups. A higher proportion of non-infected subjects was observed among wP-vaccinated participants (30.6%) compared with aP-vaccinated participants (4.3%) (Fisher’s exact test, p = 0.0194) (Fig. 1b). Conversely, the proportion of symptomatic infection was modestly higher among aP-vaccinated individuals relative to wP-vaccinated individuals (69.6% vs 47.2%, respectively) (Fisher’s exact test, p =0.1127) (Fig. 1b). Importantly, progression to symptomatic disease was associated with higher bacterial load in nasal wash, with culturable Bp detectable as early as day 3 post post-challenge and peaked around day 11 before declining following antibiotic treatment (Fig. 1c).

To identify serological features associated with protection from infection versus symptomatic disease, we performed systems serology^13^ analysis at defined time window corresponding to baseline (day −1 to day 3 post-challenge) and early post-challenge responses (day 1-7), focusing on correlates of infection and clinical disease, respectively (Fig. 1c). The baseline window, spanning day −1 to day 3 post-challenge, was selected to capture pre-existing humoral immune features present before detectable bacterial outgrowth, thereby enabling identification of antibody signatures associated with protection from infection. In contrast, the early post-challenge window, spanning days 1–7 post-challenge, was chosen to capture the initial host response following exposure but before the marked expansion in bacterial CFU, peak symptom manifestation and protocol-mandated antibiotic eradication. This window therefore allowed us to interrogate antibody features associated with modulation of clinical disease severity rather than mere correlates of exposure, high bacterial burden or treatment-induced changes. Importantly, restricting the analysis to this early interval also minimized potential confounding from azithromycin administration, which could alter bacterial persistence, antigen availability, and downstream immune maturation.

Given the presence of only a single non-infected individual among aP-vaccinated participants (Table S1), analysis of correlates of infection were restricted to the wP-vaccinated group. As an initial analysis, we examined antibody responses directed against canonical aP vaccine antigens - Fimbria 2/3 (Fim2/3), Filamentous hemagglutinin (FHA), Pertussis toxin (PT), and Pertactin- in both serum and nasal wash samples. No significant differences were observed in serum and nasal wash IgG1 responses to these antigens were observed between infected and non-infected groups (Fig. 1d-e). Comprehensive antibody profiling further demonstrated that antigen-specific IgG subclasses, IgA subclasses, IgM levels, and Fc receptor-binding capacity, and Fc-effector functions in serum and nasal wash were broadly comparable between infected and non-infected groups across all aP antigens examined (Fig. S1a). Although not statistically significant, infected individuals exhibited modestly elevated Fim2/3-specific antibody responses in serum. In nasal wash samples, infected individuals also showed trends towards higher FcγR2b and FcγR3b binding across aP antigen-specific relative to non-infected individuals (Fig. S1b). Together, these data indicate that baseline aP antigen-specific humoral responses alone are insufficient to explain resistance to Bp infection among wP-vaccinated individuals.

### Serum antibody features targeting non-canonical antigens are associated with resistance to *Bp* infection and disease

To identify previously unrecognized antibody features associated with protection, we expanded the analytical scope of our systems serology dataset to include antibody responses against a set of non-canonical aP-vaccine antigens (Table S1). These antigens were selected based on recent transposon directed insertion sequencing (TraDIS) and immunoproteomic studies implicating them in bacterial fitness, immunogenicity, and *in vivo* bacterial persistence^12^. Using this expanded antigen array, we interrogated early (day −1 to −3) humoral immune features and compared antibody landscapes between infected and non-infected individuals within the wP-primed cohort. Importantly, all antibody features were dose-corrected to control for variation in inoculum size across cohorts, reducing potential confounding and enabling unbiased identification of immune correlates. Univariate analysis revealed that resistance to Bp infection was associated with higher antibody responses targeting multiple non-canonical antigens, including FauA, TcfA, OmlA, BrkA, Yfgl, Pal, and OmpP (Fig. 2a, Table S2). These responses were characterized by increased IgA, robust Fc receptors engagement, and Fc-mediated effector functions (Fig. 2a). In contrast, infected individuals exhibited higher total IgG levels and increased FcγR2b, FcγR3a, and FcγR3b binding directed primarily toward the aP-vaccine antigen Fim2/3 (Fig. 2a, Table S2).

**Figure 2.**
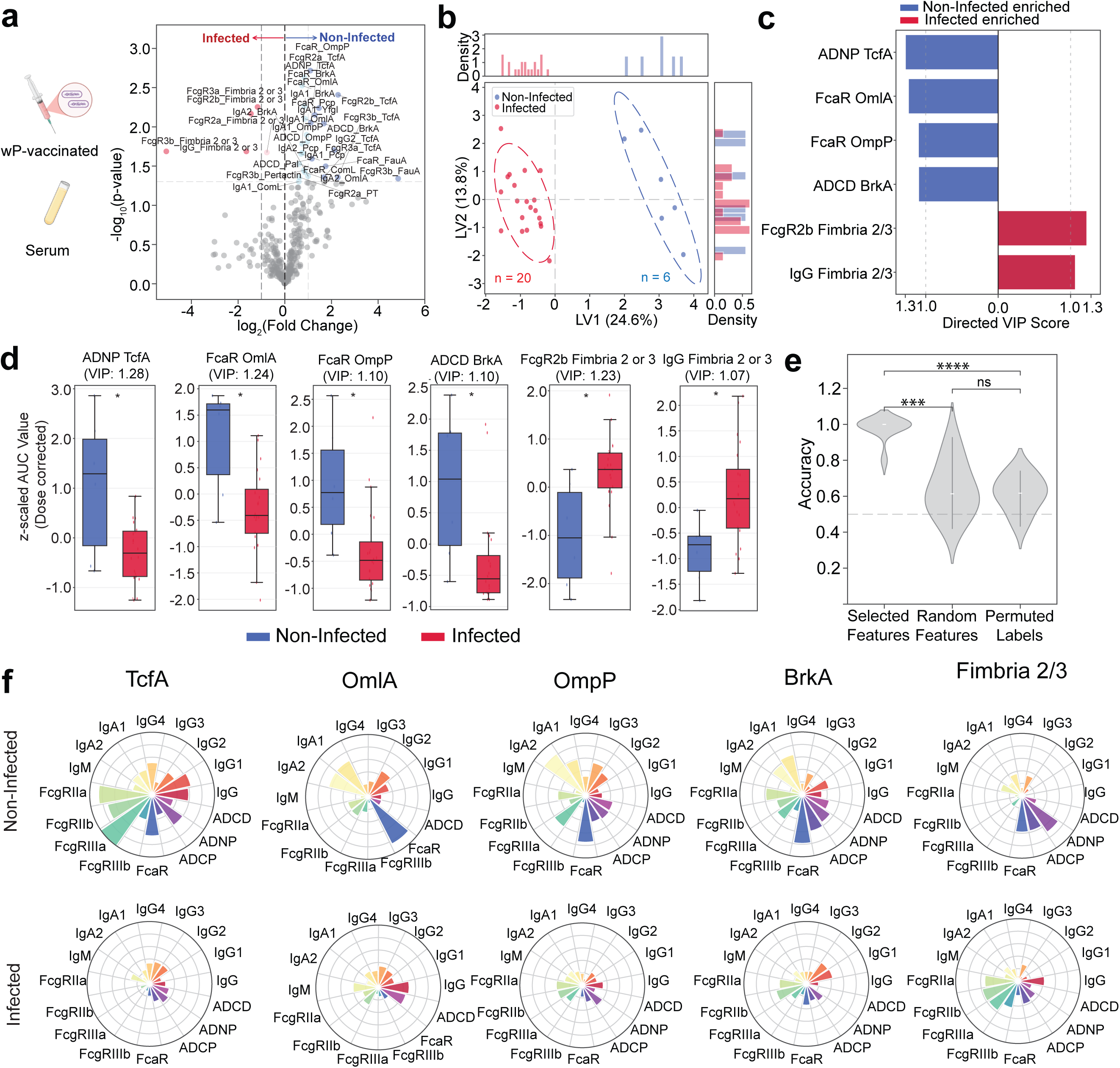
Serum antibody correlates of B. pertussis infection in wP-vaccinees. **(a)** Volcano plot showing pairwise comparison of serum antibody features between wP vaccinated non-infected (blue) and infected (red) participants following controlled human infection. Features are plotted as log₂ fold change (dose-corrected AUC day −1 to 3) versus –log₁₀ (p value). The horizontal dashed line indicates p = 0.05, and the vertical dashed line denotes the manually selected significant threshold. **(b)** Partial least squares discriminant analysis (PLS-DA) scores plot demonstrating separation between non-infected (blue) and infected (red) participants based on LASSO-selected serum antibody profiles (dose-corrected AUC day - 1 to 3). Each dot represents an individual participant; the number of individuals (n) is indicated; ellipses denote 75% confidence regions assuming a multivariate t distribution. Marginal density plots for Latent Variables- LV1 and LV2 are shown. **(c)** Directed variable importance in projection (VIP) scores highlighting top discriminative serum features (VIP > 1.0) enriched in non-infected (blue) or infected (red) individuals. (d) Boxplots representing medians, interquartile ranges [IQRs], minima, and maxima of representative discriminative features (VIP >1.0, panel c) comparing non-infected (blue) and infected (red) individuals; the number of individuals is the same as in panel **(b)**. The measurements were dose-corrected and z-scored for AUC day −1 to 3. Significances were assessed using Mann–Whitney U tests and corrected for multiple testing using the Benjamini–Hochberg procedure. Asterisks indicate adjusted P values with *P < 0.05, **P < 0.01, ***P < 0.001. (e) The model performance was validated using permutation tests. Classification accuracy is shown for models built using selected features (panel c), random features, or permuted labels. Violin plots show the distribution from 100 repetitions for each model type. P values represent the median exact permutation-derived P value across repetitions. Significances were assessed using Mann–Whitney U tests and corrected for multiple testing using the Benjamini–Hochberg procedure. Asterisks indicate adjusted P values with *P < 0.05, **P < 0.01, ***P < 0.001. (f) Radar/Polar plots depicting mean percentile ranks of antigen-specific antibody feature distributions for representative discriminative antigens (VIP >1.0, panel c) in non-infected (top) versus infected (bottom) individuals. Percentile rank scores were determined for each antibody feature across all individuals.

To integrate these features and identify multivariate correlates of protection, we applied supervised Least Absolute Shrinkage and Selection Operator (LASSO)-regularized partial least squares discriminant analysis (PLS-DA). This model robustly separated infected and non-infected individuals (Fig. 2b) and identified a concise set of discriminatory antibody features (Variable Importance in projection, VIP > 1) driving group separation (Fig. 2c). Protective signatures in non-infected individuals were dominated by FcαR-binding antibodies to OmlA and OmpP, along with antibody-dependent neutrophil phagocytosis (ADNP) and complement deposition (ADCD) targeting TcfA and BrkA, respectively. Conversely, infected subjects were enriched for IgG and FcγR2b-binding antibodies directed against Fim2/3 (Fig. 2c, d). High-importance features (VIP>1) displayed higher-magnitude distributions within their respective outcome group at univariate level (Fig. 2d). Cross-validation demonstrated near-perfect classification accuracy (∼1.0), significantly exceeding random or permuted models (Fig. 2e). The LASSO algorithm selects a minimal non-redundant set of features that drives overall variation across groups, while excluding features that correlate highly with those that are selected. Thus, to fully capture the spectrum of antibody features that differentiate the two groups, correlation networks were generated across the non-infected group for the LASSO-selected features (Table S3). Finally, we compared the qualitative and quantitative architecture of the humoral response elicited by antigens represented among the highest-weighted VIP antibody features across infected and non-infected wP-recipients. Non-infected subjects exhibited broader, higher-magnitude, and more polyfunctional antibody responses across multiple IgG and IgA subclasses, coupled with stronger antibody-dependent cellular phagocytosis (ADCP), ADNP, and ADCD activity against TcfA, OmlA, OmpP, and BrkA (Fig. 2f). In contrast, infected subjects displayed mounted lower-magnitude responses across these antigens, with reduced functional activity, with notable exception of Fim2/3 (Fig. 2f). Together, these data reveal distinct humoral immune architectures associated with resistance or susceptibility to Bp infection among wP-vaccinated participants.

We next examined serological correlates of symptomatology (days 1-7) clinical disease by uni- and multivariate analysis of serum samples collected during the early post-challenge windows. (days 1-7). Among aP-vaccinated participants, asymptomatic individuals displayed broader and more robust humoral responses across several Bp antigens, including OmlA, Pertactin, FauA, and BP1485 (Fig. S2a, Table S2). In comparison, none of the antibody features were significantly enriched in symptomatic participants. PLS-DA modeling identified discriminative features (VIP>1) that reliably and with high classification accuracy separated asymptomatic and symptomatic groups (Fig. S2b, c), with asymptomatic subjects displaying higher-magnitude IgG- and IgG3- directed against OmlA, FcgR2b-binding capacity of BP1485-specific antibodies, and IgG-FauA (Fig. S2c, d, e, f, Table S3). Similarly, the wP-vaccinated cohort yielded distinct immune correlates of symptomatology. Asymptomatic individuals showed higher levels of pertussis toxin (PT)-IgG1, OmlA-IgG1, and FcαR engagement by OmpP-specific antibodies, symptomatic subjects showed elevated responses towards FcαR engagement by YfgL-specific antibodies and IgA2-Pcp (Fig. S3a, Table S2, Fig. S3b-d, f). These features yielded a PLS-DA model with strong classification performance (Fig. S3b-e, Table S3).

Collectively, these findings demonstrate that serum antibody responses targeting a set of non-canonical Bp antigens are strongly associated with resistance to infection and reduced symptomatology. Moreover, the immune signatures associated with infection and symptom development differ and are shaped by prior vaccination history, underscoring the importance of antigenic breadth and Fc-functional quality in protective anti-pertussis immunity.

### Distinct mucosal antibody signatures are linked to resistance against Bp infection and disease development

Given that Bp is a respiratory pathogen that primarily colonizes the posterior nasopharynx, we next investigated mucosal antibody profiles from nasal wash samples to define mucosal correlates of resistance to infection and symptomatology. Baseline (Day −1 to 3) profiling of nasal wash samples from wP-vaccinated participants revealed that non-infected individuals mounted higher-magnitude of IgG1 and IgG3 targeting FauA, ACT, and pertactin, along with engagement of FcγR2a and FcγR3a by antibodies targeting BP1485, OmpA, TcfA and BP0183 (Fig. 3a, Table S2). No features were found to be significantly higher in the infected participants. Supervised LASSO-PLSDA robustly separated non-infected and infected individuals (Fig. 3b) and identified a discrete set of discriminatory mucosal antibody features (VIP > 1) (Fig. 3b-c). Non-infected individuals were enriched for FHA- and TcfA-specific IgA1, Pertactin-specific IgG3, FauA- specific IgG1, and TcfA- and OmlA- specific IgM, whereas infected individuals showed enrichment of FcαR-binding antibodies targeting TamB and YfgL (Fig. 3c). Although univariate analysis indicated that none of the individual VIP features reached statistical significance, the multivariate model achieved high classification accuracy (∼1), indicating that the combined antibody features robustly distinguish infected from non-infected individuals (Fig. 3e). Co-correlate analysis revealed that LASSO-selected features formed dense intramucosal correlation networks within the nasal wash compartment (Table S4). Architectural analysis of LASSO-selected antigens revealed that non-infected individuals mounted broader and more diverse mucosal antibody profiles spanning multiple IgG and IgA subclasses, and Fc receptor interactions. Conversely, infected individuals exhibited narrower and lower-magnitude mucosal responses with limited Fc engagement (Fig. 3f).

**Figure 3.**
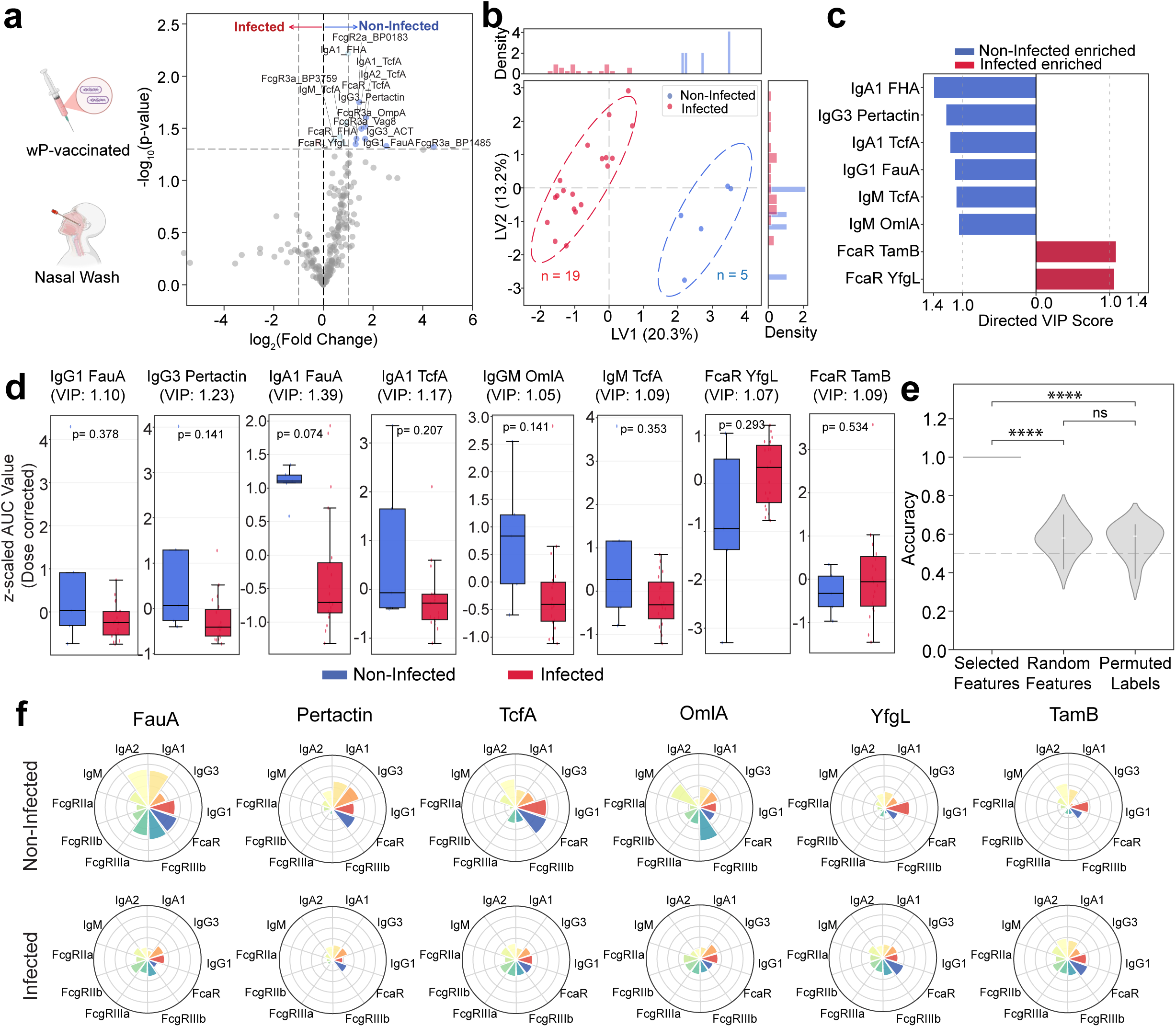
Mucosal antibody correlates of B. pertussis infection in wP-vaccinees. **(a)** Volcano plot showing pairwise comparison of nasal wash antibody features between wP vaccinated non-infected (blue) and infected (red) participants following controlled human infection. Features are plotted as log₂ fold change (dose-corrected AUC day −1 to 3) versus –log₁₀ (p value). The horizontal dashed line indicates p = 0.05, and the vertical dashed line denotes the manually selected significant threshold. **(b)** PLS-DA scores plot demonstrating separation between non-infected (blue) and infected (red) participants based on LASSO-selected nasal wash antibody profiles (dose-corrected AUC day −1 to 3). Each dot represents an individual participant; the number of individuals (n) is indicated; ellipses denote 75% confidence regions assuming a multivariate t distribution. Marginal density plots for LV1 and LV2 are shown. **(c)** Directed VIP scores highlighting top discriminative nasal wash features (VIP > 1.0) enriched in non-infected (blue) or infected (red) individuals. (d) Boxplots representing medians, IQRs, minima, and maxima of representative discriminative features (VIP >1.0, panel c) comparing non-infected (blue) and infected (red) individuals; the number of individuals is the same as in panel **(b)**. The measurements were dose-corrected and z-scored for AUC day −1 to 3. Significances were assessed using Mann–Whitney U tests and corrected for multiple testing using the Benjamini–Hochberg procedure. Asterisks indicate adjusted P values with *P < 0.05, **P < 0.01, ***P < 0.001. (e) The model performance was validated using permutation tests. Classification accuracy is shown for models built using selected features (panel c), random features, or permuted labels. Violin plots show the distribution from 100 repetitions for each model type. P values represent the median exact permutation-derived P value across repetitions. Significances were assessed using Mann–Whitney U tests and corrected for multiple testing using the Benjamini–Hochberg procedure. Asterisks indicate adjusted P values with *P < 0.05, **P < 0.01, ***P < 0.001. (f) Radar/Polar plots depicting mean percentile ranks of antigen-specific antibody feature distributions for representative discriminative antigens (VIP >1.0, panel c) in non-infected (top) versus infected (bottom) individuals. Percentile rank scores were determined for each antibody feature across all individuals.

We next evaluated early mucosal correlates of symptomatology (days 1–7) in aP and wP participants (Figures S4 and S5). Asymptomatic aP-primed subjects exclusively displayed an expanded repertoire of FcγR3a against PT and pertactin, and FcγR3b binding IgG against pertactin, OmlA, and BP3759 (Fig. S4a, Table S2). Subsequently, LASSO/PLS-DA defined unique features between the asymptomatic and symptomatic groups. The most discriminatory features included FcγR3b and FcαR binding capacity of OmlA- and vag8-specific antibodies, respectively for asymptomatic participants, in the case of asymptomatic subjects and FcγR2b and FcγR3b binding IgG against Pcp in symptomatic subjects (Fig. S4b-e, Table S4). Further analysis revealed that asymptomatic subjects produced a robust polyfunctional antibody response against key antigens (VIP >1), in stark contrast to symptomatic subjects (Fig. S4f). At univariate level, wP-primed asymptomatic participants showed elevated BamA-specific IgG1 and enhanced FcγR3a binding capacity of PT-specific antibodies in the nasal mucosa, whereas no individual features were significantly enriched in the symptomatic participants (Fig S5a, Table S2). Supervised LASSO-PLSDA further identified a discrete set of discriminatory mucosal features (VIP > 1), including: IgG1 BamA, IgA2 BP3760, IgA2 Vag8, and FcαR-binding Vag8-specific antibodies, that collectively separated asymptomatic from symptomatic individuals (Fig. S5b–c, Table S4). Although these individual features did not reach statistical significance in univariate testing (Fig. S5d), their combined integration yielded a model with significantly higher predictive accuracy compared to random feature selection or permuted labels (∼1.0 accuracy) (Fig. S5e). Architectural analysis of the LASSO-selected antigens revealed that asymptomatic subjects mounted a broader and more diversified subclass responses spanning IgG1, IgA, and Fc receptor interactions across BamA, BP3760, and Vag8, than symptomatic participants (Fig. S5f). In contrast, symptomatic individuals demonstrated narrower and lower-magnitude Fc engagement profiles against the same antigen set.

Collectively, these findings indicate that resistance to Bp infection and symptom development is associated with stronger and broader IgA-dominated mucosal antibody responses targeting select non-canonical Bp antigens. Notably, the antigenic determinants driving mucosal discrimination were largely distinct from those identified in serum, suggesting compartmentalized immune correlates of protection.

### Protection from pertussis infection and clinical manifestation requires coordination between systemic and mucosal antibody responses

Effective protection against respiratory pathogens likely requires coordination between circulating and mucosal antibody responses^15–17^, therefore, we integrated serum and nasal wash antibody features to define compartment-spanning correlates of resistance to infection in wP-vaccinated individuals (Figure 4). Univariate analysis of the combined datasets identified distinct systemic and mucosal antibody signatures associated with resistance to infection. The antibody signatures in serum in non-infected individuals were dominated by IgG responses to TcfA, FcγR-engaging TcfA-specific antibodies, ADNP activity directed against TcfA, and complement-activating readouts targeting BrkA, Pal and OmpP among other antibody correlates. In contrast, the nasal wash compartment was enriched for mucosal isotypes and FcαR linked correlates, including IgA1-, IgA2- and FcαR-binding TcfA-specific antibodies, FcγR-binding responses to OmpA and BP0183, and IgG responses to ACT, FauA and Pertactin. Notably, TcfA-specific responses in were represented in both compartments, highlighting TcfA as a shared systemic and mucosal correlate of infection resistance (Fig. 4a, Table S2). In contrast, infected individuals exhibited enrichment of only serum IgG levels and Fc-binding capacity directed toward Fim2/3 (Fig. 4a, Table S2).

**Figure 4.**
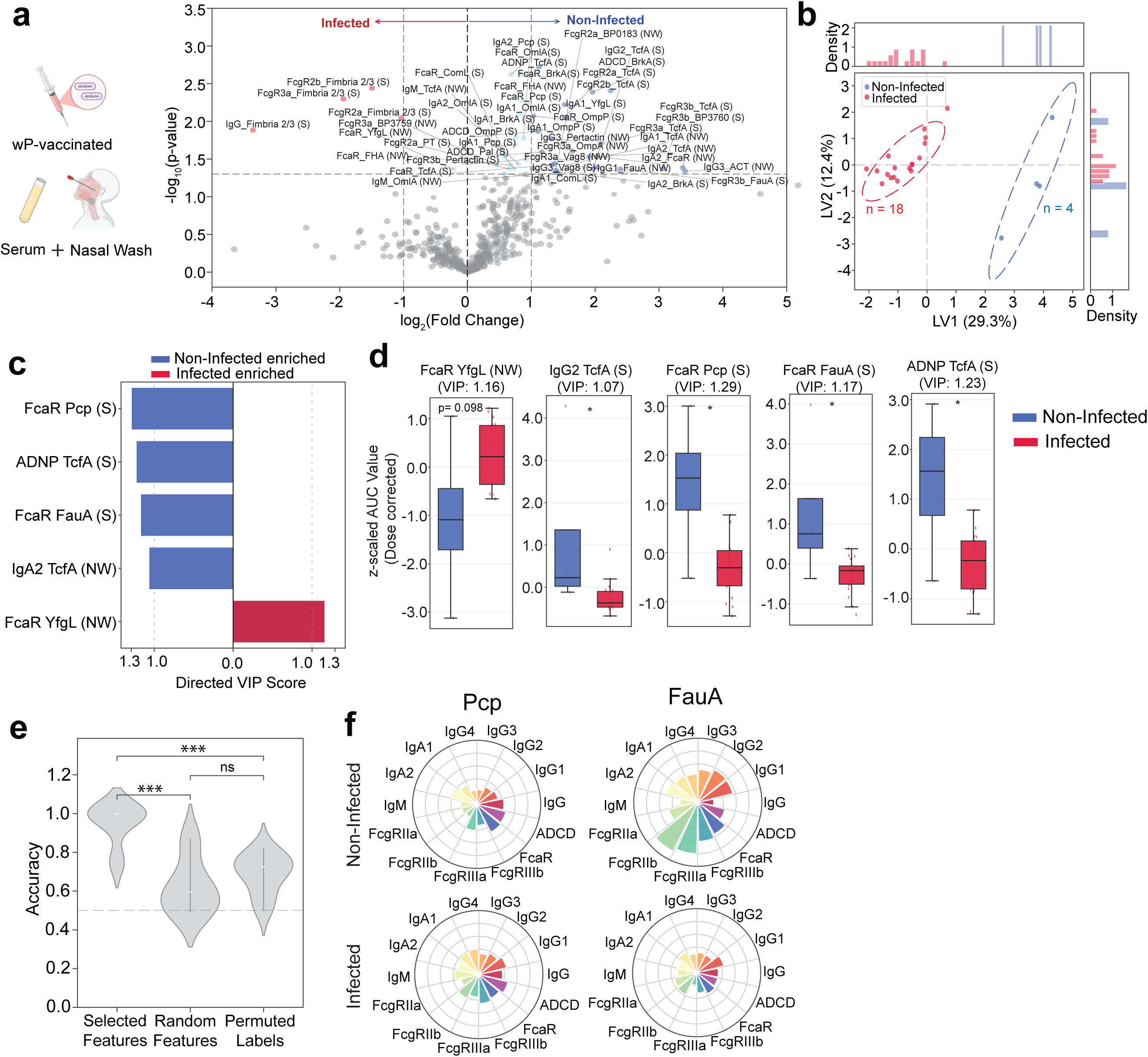
Serum and mucosal antibody correlates of B. pertussis infection in wP-vaccinees. **(a)** Volcano plot showing pairwise comparison of combined serum (S) and nasal wash (NW) antibody features between wP vaccinated non-infected (blue) and infected (red) participants following controlled human infection. Features are plotted as log₂ fold change (dose-corrected AUC day −1 to 3) versus –log₁₀ (p value). The horizontal dashed line indicates p = 0.05, and the vertical dashed line denotes the manually selected significant threshold. **(b)** PLS-DA scores plot demonstrating separation between non-infected (blue) and infected (red) participants based on LASSO-selected serum and nasal wash antibody features (dose-corrected AUC day −1 to 3). Each dot represents an individual participant; the number of individuals (n) is indicated; ellipses denote 75% confidence regions assuming a multivariate t distribution. Marginal density plots for LV1 and LV2 are shown. **(c)** Directed VIP scores highlighting top discriminative antibody features (VIP > 1.0) enriched in non-infected (blue) or infected (red) individuals. (d) Boxplots representing medians, IQRs, minima, and maxima of representative discriminative features (VIP >1.0, panel c) comparing non-infected (blue) and infected (red) individuals; the number of individuals is the same as in panel **(b)**. The measurements were dose-corrected and z-scored for AUC day −1 to 3. Significances were assessed using Mann–Whitney U tests and corrected for multiple testing using the Benjamini–Hochberg procedure. Asterisks indicate adjusted P values with *P < 0.05, **P < 0.01, ***P < 0.001. **(e)** The model performance was validated using permutation tests. Classification accuracy is shown for models built using selected features (panel c), random features, or permuted labels. Violin plots show the distribution from 100 repetitions for each model type. P values represent the median exact permutation-derived P value across repetitions. Significances were assessed using Mann–Whitney U tests and corrected for multiple testing using the Benjamini–Hochberg procedure. Asterisks indicate adjusted P values with *P < 0.05, **P < 0.01, ***P < 0.001. (f) Radar/Polar plots depicting mean percentile ranks of antigen-specific antibody feature distributions for representative discriminative antigens (VIP >1.0, panel c) in non-infected (top) versus infected (bottom) individuals. Radar plots for TcfA and YfgL are given in Fig. 2f & 3f, respectively. Percentile rank scores were determined for each antibody feature across all individuals.

Supervised LASSO-PLS-DA analysis robustly separated non-infected and infected subjects (Fig. 4b), identifying a discrete panel of discriminatory features (VIP > 1) spanning both systemic and mucosal compartments. Non-infected individuals were enriched for serum FcαR-binding antibodies targeting Pcp and FauA, serum ADNP activity directed against TcfA, and IgA2-TcfA responses in nasal wash (Fig. 4b-d). In contrast, infected individuals were enriched for FcαR-binding YfgL-specific antibodies in nasal wash samples (Fig. 4b-d). Although individual features showed only modest statistical significance (Fig. 4d), integration of the selected features yielded significantly improved classification accuracy compared to random or permuted models (Fig. 4e). Architectural analysis further revealed that non-infected individuals mounted broader subclass diversification and enhanced Fc receptor engagement against Pcp and FauA, whereas infected subjects displayed comparatively narrower functional profiles (Fig. 4f). Together, these data indicate that resistance to infection in wP-vaccinated individuals is associated with antibody responses spanning both serum and mucosal compartments.

Next, using the integrated systemic and mucosal dataset from days 1-7 post-challenge, we examined correlates of symptomatology. Among aP-primed individuals, asymptomatic subjects exhibited enrichment predominantly within the serum compartment, characterized by elevated IgG subclasses and enhanced FcγR-binding capacity directed against OmlA, BP1485, FauA, and TcfA (FigS6a, Table S2). Multivariate analysis identified top differentiating features in asymptomatic subjects (Fig. S6b-c). While individual univariate comparisons of LASSO-selected features (VIP>1) were modest (Fig. S6d), multivariate integration yielded significantly improved predictive accuracy compared to random feature selection or permuted labels (Fig. S6e). Furthermore, correlation network analysis revealed that LASSO-selected features in non-infected individuals were highly interconnected with both serum and mucosal antibody responses, forming coordinated cross-compartment humoral networks (Table S5).

Similarly, in wP-primed subjects, serum correlates predominated over mucosal features in distinguishing asymptomatic from symptomatic individuals. Asymptomatic subjects exhibited elevated serum IgG1 levels against BamA, OmlA and PT along with enhanced FcγR engaging antibodies targeting OmlA and Pcp, and enrichment of IgG1-BamA in nasal wash (Fig. S7a, Table S2). Multivariate modeling robustly separated the two groups, identifying IgG1-BamA in nasal wash compartment, serum IgG4- and IgG2-OmlA and serum IgG3-Pertactin as the immune correlates enriched in asymptomatic group (Fig. S7c, Table S5). Although none of these individual features reached statistical significance in isolation (Fig. S7d), the integrated model achieved significantly higher classification accuracy compared to random or permuted controls (Fig. S7e). In line with other models, architecture analysis demonstrated that asymptomatic subjects exhibited broader subclass diversification and enhanced Fc receptor engagement across OmlA, BamA and Pertactin, while symptomatic individuals displayed comparatively restricted functional profiles (Fig. S7f, S5f, S3f). Although LASSO-selected discriminatory features were compartment-skewed in individual models, correlate network analysis (Table S5) revealed that these features were highly interconnected with nasal wash antibody responses across all three models, indicating functional cross-compartment coordination.

Collectively, these findings demonstrate that resistance to infection and clinical disease is not confined to a single anatomical compartment but instead reflects coordinated systemic–mucosal humoral immune architectures.

### LASSO-selected antibody features outperform canonical vaccine antigens in predicting resistance to infection

Subsequently, to examine the overlap among the LASSO-selected VIP (>1) features identified in the serum-only (Fig. 2), nasal mucosa–only (Fig. 3), and combined (serum + nasal mucosa, Fig. 4) compartments, we performed an UpSet plot analysis, which revealed the magnitude of overlap between these compartments. Intersection analysis of correlates in wP-vaccinated participants revealed both shared and compartment-specific antibody features, highlighting partially overlapping yet distinct humoral architectures underlying resistance to infection (Fig. 5a, Table S6).

**Figure 5.**
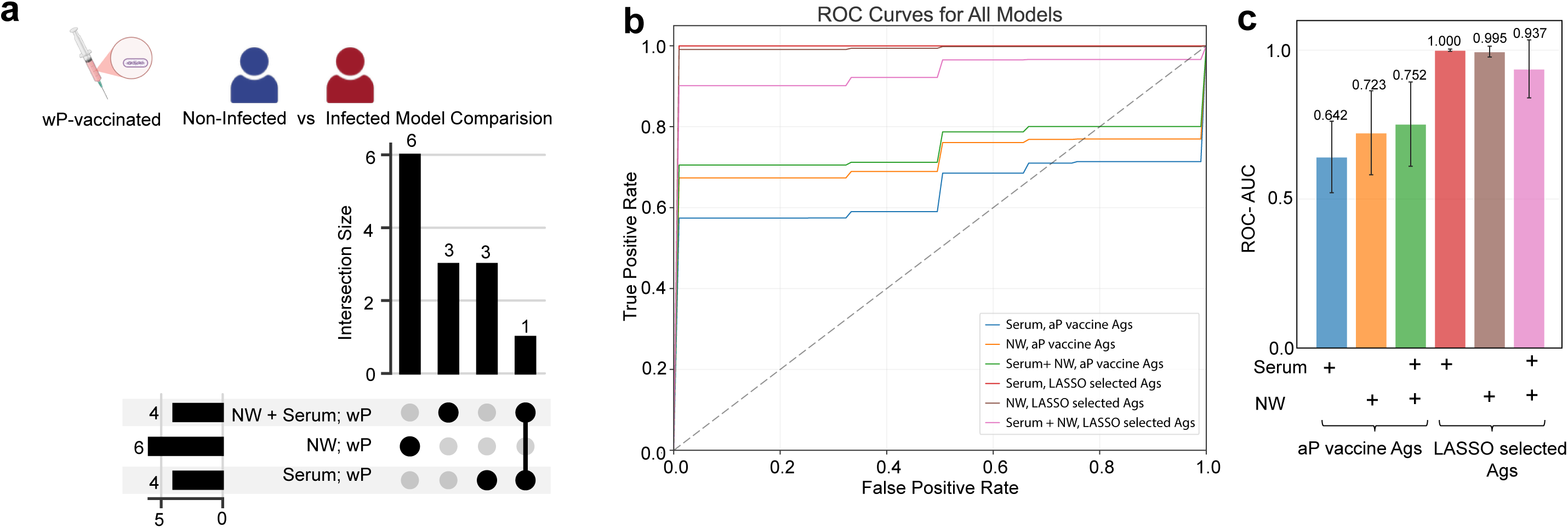
LASSO-selected antibody features outperform classification of B. pertussis infection in wP vaccinees. (a) UpSet plot showing the overlap of discriminative antigen sets derived from LASSO/PLSDA models using antibody features in serum, nasal wash, or combined serum + nasal wash in wP-vaccinated participants. (b) Receiver operating characteristic (ROC) curves comparing model performance for classifying non-infected versus infected individuals using serum alone, nasal wash alone, or combined serum + nasal wash antibody features across antigen classes (aP vaccine antigens, LASSO-selected antigens, and microbiome antigens). (c) Bar plot summarizing ROC–AUC values for each model. Bars represent mean ROC–AUC ± S.D.

Next, we evaluated the predictive capacity of compartment-specific and integrated models using ROC analysis (Fig. 5b). Models constructed using canonical aP vaccine antigens demonstrated only modest discriminatory power, with AUC values ranging from 0.642 (serum) to 0.752 (serum + nasal wash) (Fig. 5c). In contrast, models built using LASSO-selected features markedly improved classification performance. Serum-only and nasal wash-only LASSO models achieved near-perfect discrimination (AUC = 1.000 and 0.995, respectively), while the integrated serum + nasal wash model maintained high predictive accuracy (AUC = 0.937) (Fig. 5b–c). These findings demonstrate that feature selection using LASSO-PLS-DA substantially improves predictive performance beyond that of canonical vaccine antigen–based models.

To determine whether similar compartmental relationships were observed in clinical disease models, we performed parallel intersection and ROC analyses in aP- and wP- vaccinated cohorts (Fig. S8). In the wP-vaccinated cohort, minimal overlap was observed between integrated (serum + nasal wash) and nasal wash-only correlates, with only one out of four shared features. In contrast, the aP-vaccinated cohort demonstrated greater overlap between integrated and serum-derived correlates, with three shared features and one feature common to both the integrated and nasal wash compartments (Fig. S8a). Notably, one nasal wash correlate was common between wP- and aP-vaccinated subjects (Table S7).

Consistent with infection analysis, predictive performance of symptomatology in both vaccine groups improved substantially when using LASSO-selected antigens (Fig. S8b-e). Integrated systemic–mucosal models achieved near-perfect discrimination in both aP- and wP-vaccinated cohorts (AUC ∼1.0), whereas compartment-specific LASSO models yielded moderate-to-high performance (AUC ∼0.7–0.9). In contrast, models based solely on canonical aP vaccine antigens demonstrated limited predictive capacity (Fig. S8b–e). Collectively, these findings demonstrate that optimized Fc-effector feature selection markedly improves classification of both resistance to infection and symptom outcomes and that coordinated cross-compartment antibody architectures underpin robust prediction of pertussis disease phenotypes.

## Discussion

The inability of aP vaccines to prevent colonization and onward transmission remains a major public health concern. A central barrier to rational next-generation vaccine design has been the absence of well-defined immune correlates of protection for pertussis. Unlike diseases such as tetanus or measles, where defined antibody thresholds predict immunity, no antibody level or biomarker has been universally established as a correlate of protection against pertussis infection or disease^18^. Historically, serum anti–PT IgG has been considered a potential surrogate marker, with some studies suggesting that anti-PT IgG >5 IU/mL is associated with reduced risk of symptomatic disease^18^. However, these benchmarks are imprecise and primarily reflect protection against severe clinical manifestations rather than prevention of infection. The complexity of pertussis immunity encompassing toxin-neutralizing antibodies, opsonophagocytic antibodies, mucosal IgA, and Th1/Th17-driven cellular responses^10,19–24^ has contributed to uncertainty regarding the precise immunological mechanisms and antigenic targets required for complete protective immunity. To address this gap and identify yet uncharacterized correlates of protection, we leveraged the first North American controlled human infection model (CHIM) cohort, profiling longitudinal systemic and mucosal antibody responses to an expanded panel of *Bordetella pertussis* antigens in 59 participants.

Baseline responses to canonical aP antigens (PT, FHA, pertactin, and Fim2/3) did not discriminate infection outcomes, aligning with epidemiologic and experimental evidence that aP-induced immunity protects against severe disease but incompletely prevents infection or transmission^3,25,26^. In contrast, resistance to infection in wP-vaccinated individuals was associated with antibody responses targeting non-canonical antigens previously identified using convalescent baboon sera^12^. The broader antigenic recognition observed in wP-primed individuals likely reflects the epitope diversity inherent to whole-cell formulations, potentially facilitating more effective opsonophagocytic and complement-mediated clearance. Several of the identified antigens have well-established roles in pathogenesis. TcfA, a tracheal colonization factor, contributes to adherence and airway persistence in murine models and has been exploited as a diagnostic biomarker due to its specificity^27^. BrkA and Vag8 function as complement-evasion proteins that inhibit classical and alternative pathway activation^28,29^. Outer membrane-associated proteins including OmlA, OmpP, Pal, and BamA/YfgL (BamB) are involved in membrane integrity and biogenesis^12^, while FauA mediates iron acquisition, a critical determinant of *in vivo* fitness^30^. Antibody responses directed against these targets may enhance complement deposition (ADCD), neutrophil phagocytosis (ADNP), and other Fc-receptor–dependent effector functions, collectively limiting bacterial establishment at mucosal surfaces. Together, these findings underscore that qualitative features and antigenic breadth, rather than antibody magnitude alone—are central to protection. Convergence between CHIM-derived signatures and antigens prioritized in baboon studies ^12^ further supports their biological relevance.

Systemic and mucosal antibody responses following exposure were strongly shaped by early-life vaccine priming history and were associated with differential susceptibility to infection. wP-primed participants demonstrated significantly greater resistance to infection, whereas aP-primed individuals exhibited higher bacterial burdens and more frequent establishment of infection. High-dimensional systems serology revealed that infection resistance was not explained by responses to conventional DTaP antigens alone, but instead associated with polyfunctional, Fc-engaging antibodies targeting a broader antigenic repertoire. These correlates were distributed across systemic and mucosal compartments and differed by priming history, underscoring that sterilizing immunity reflects coordinated, multi-antigen humoral responses rather than antibody magnitude to canonical vaccine components.

In contrast to resistance to infection, mitigation of clinical symptoms emerged as a partially independent immunologic outcome. Reduction in disease severity was observed in both vaccine groups and associated with vaccine history–specific antibody signatures rather than complete bacterial clearance. In aP-primed individuals, symptom protection correlated predominantly with systemic IgG subclass and Fc receptor–binding responses to OmlA, BP1485, FauA, and TcfA. In wP-primed individuals, overlapping but distinct features included responses to BamA, OmlA, PT, and pertactin. These findings suggest that antibody-mediated modulation of toxin activity and inflammatory pathways can attenuate clinical manifestations even when infection persists. Correlation network analyses further demonstrated that these discriminatory features, although compartment-skewed in multivariate models, were embedded within coordinated systemic–mucosal humoral networks, supporting the concept that integrated cross-compartment immunity contributes to optimal disease control.

Expansion of the antigen panel to include recently identified TraDIS and immunoproteomic hits^12^, enabled identification of a concise set of antibody features that robustly predicted resistance to infection in wP-primed adults. Integrated serum and nasal analyses highlighted TcfA as a shared correlate across compartments, while mucosal features such as IgG1-BamA and FcγR3a-binding PT distinguished asymptomatic wP-primed individuals. These findings suggest that aP priming preferentially elicits systemic correlates of disease attenuation, whereas wP priming supports broader immunity encompassing functionally relevant mucosal responses.

This study has limitations. The CHIM was conducted in a relatively small cohort of healthy adults (n=59) with documented childhood vaccination histories, which may limit extrapolation to infants—who bear the highest disease burden and have distinct immune ontogeny—or to older adults with waning immunity. In addition, CHIM infection dynamics may differ in magnitude, duration, and immune activation kinetics from natural community-acquired infections, where exposure dose, route, timing, co-infections, and host background are more heterogeneous. Therefore, although the models provide important mechanistic insights, its predictive power and broader applicability would be strengthened by validation in larger, independent cohorts that include more diverse age groups, vaccination backgrounds, exposure settings, and infection outcomes.

Collectively, these findings provide a mechanistic framework to guide next-generation pertussis vaccine development. Identification of candidate antigens, including TcfA, FauA, OmlA, OmpP, BrkA, Pal, BamA, and Vag8, supports strategies that expand antigenic breadth and enhance functional antibody responses, with the goal of achieving more effective systemic and mucosal protection against this resurgent pathogen.

**Figure S1.**
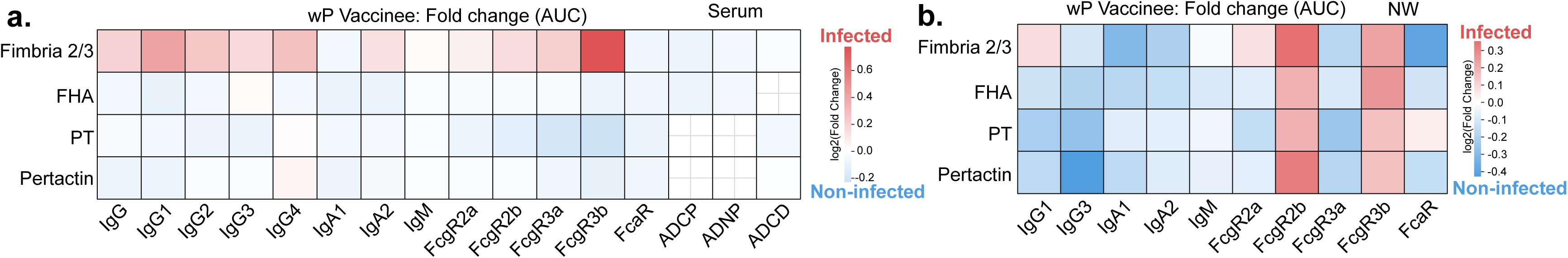
Antibody profiles of wP vaccinees. (a-b) Heatmap demonstrating log2 fold changes in aP-antigen specific antibody features (AUC, day −1 to 3) between infected and non-infected participants in serum (a) and nasal wash (b). Color intensity reflects the direction and magnitude of enrichment, with red indicating features elevated in infected individuals and blue indicating features enriched in non-infected individuals. Features not examined are crossed. Significances were assessed using Mann–Whitney U tests and corrected for multiple testing using the Benjamini–Hochberg procedure. Asterisks indicate adjusted P values with *P < 0.05, **P < 0.01, ***P < 0.001

**Figure S2.**
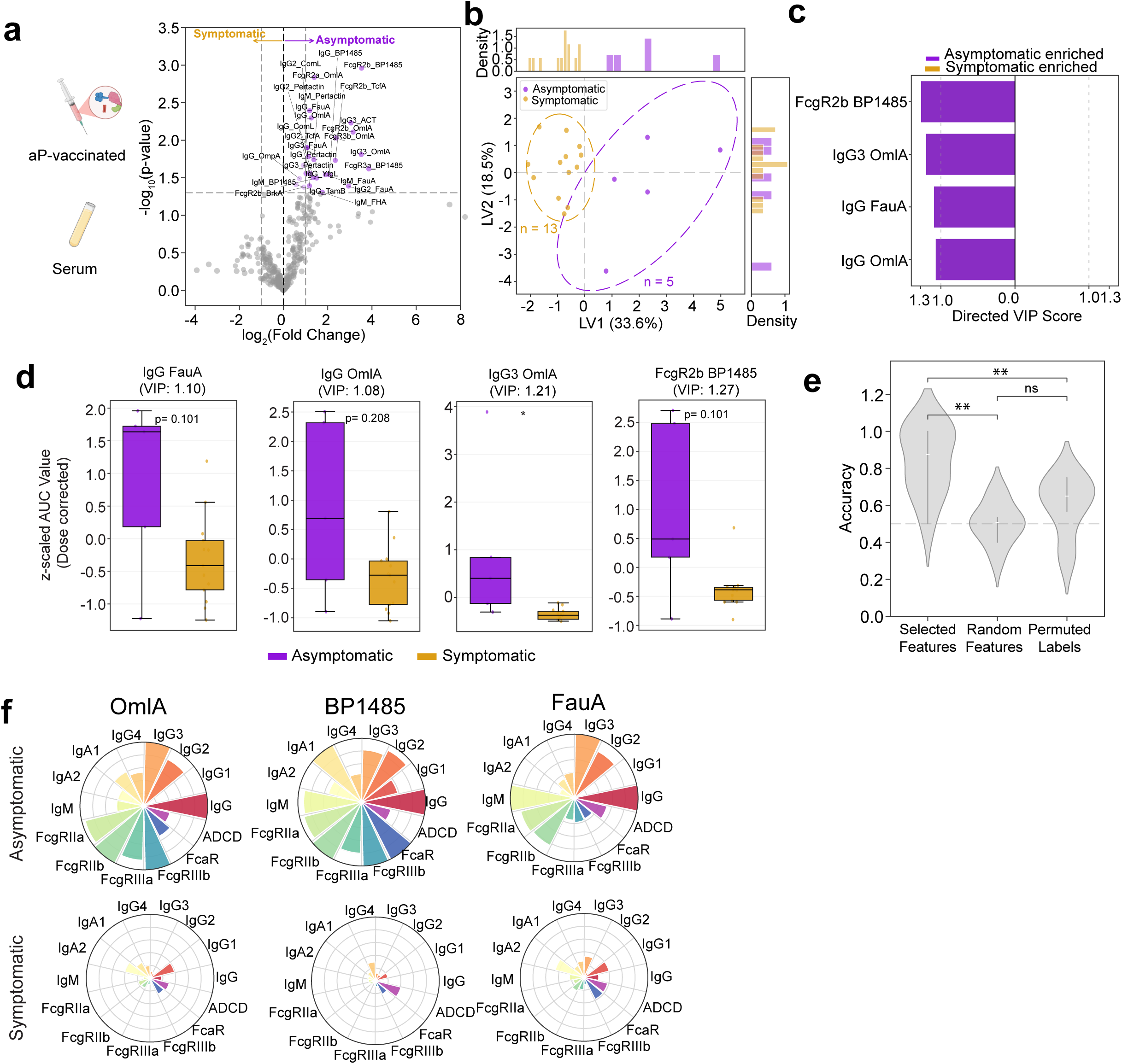
Serum antibody correlates of B. pertussis symptomology in aP-vaccinees. **(a)** Volcano plot showing pairwise comparison depicting serum antibody features between aP vaccinated asymptomatic (purple) and symptomatic (yellow) participants following controlled human infection. Features are plotted as log₂ fold change (dose-corrected AUC day 1 to 7) versus –log₁₀ (p value). The horizontal dashed line indicates p = 0.05, and the vertical dashed line denotes the manually selected significant threshold. **(b)** Partial least squares discriminant analysis (PLS-DA) scores plot demonstrating separation between asymptomatic (purple) or symptomatic (yellow) participants based on LASSO-selected serum antibody profiles (dose-corrected AUC day 1 to 7). Each dot represents an individual participant; the number of individuals (n) is indicated; ellipses denote 75% confidence regions assuming a multivariate t distribution. Marginal density plots for Latent Variables- LV1 and LV2 are shown. **(c)** Directed variable importance in projection (VIP) scores highlighting top discriminatory serum features (VIP > 1.0) enriched in asymptomatic (purple) or symptomatic (yellow) individuals. (d) Boxplots representing medians, interquartile ranges [IQRs], minima, and maxima of representative discriminative features (VIP >1.0, panel c) comparing asymptomatic (purple) or symptomatic (yellow) individuals; the number of individuals is the same as in panel **(b)**. The measurements were dose-corrected and z-scored for AUC day 1 to 7. Significances were assessed using Mann–Whitney U tests and corrected for multiple testing using the Benjamini–Hochberg procedure. Asterisks indicate adjusted P values with *P < 0.05, **P < 0.01, ***P < 0.001. (e) The model performance was validated using permutation tests. Classification accuracy is shown for models built using selected features (panel c), random features, or permuted labels. Violin plots show the distribution from 100 repetitions for each model type. P values represent the median exact permutation-derived P value across repetitions. Significances were assessed using Mann–Whitney U tests and corrected for multiple testing using the Benjamini–Hochberg procedure. Asterisks indicate adjusted P values with *P < 0.05, **P < 0.01, ***P < 0.001. (f) Radar/Polar plots depicting mean percentile ranks of antigen-specific antibody feature distributions for representative discriminative antigens (VIP >1.0, panel c) in non-infected (top) versus infected (bottom) individuals. Percentile rank scores were determined for each antibody feature across all individuals.

**Figure S3.**
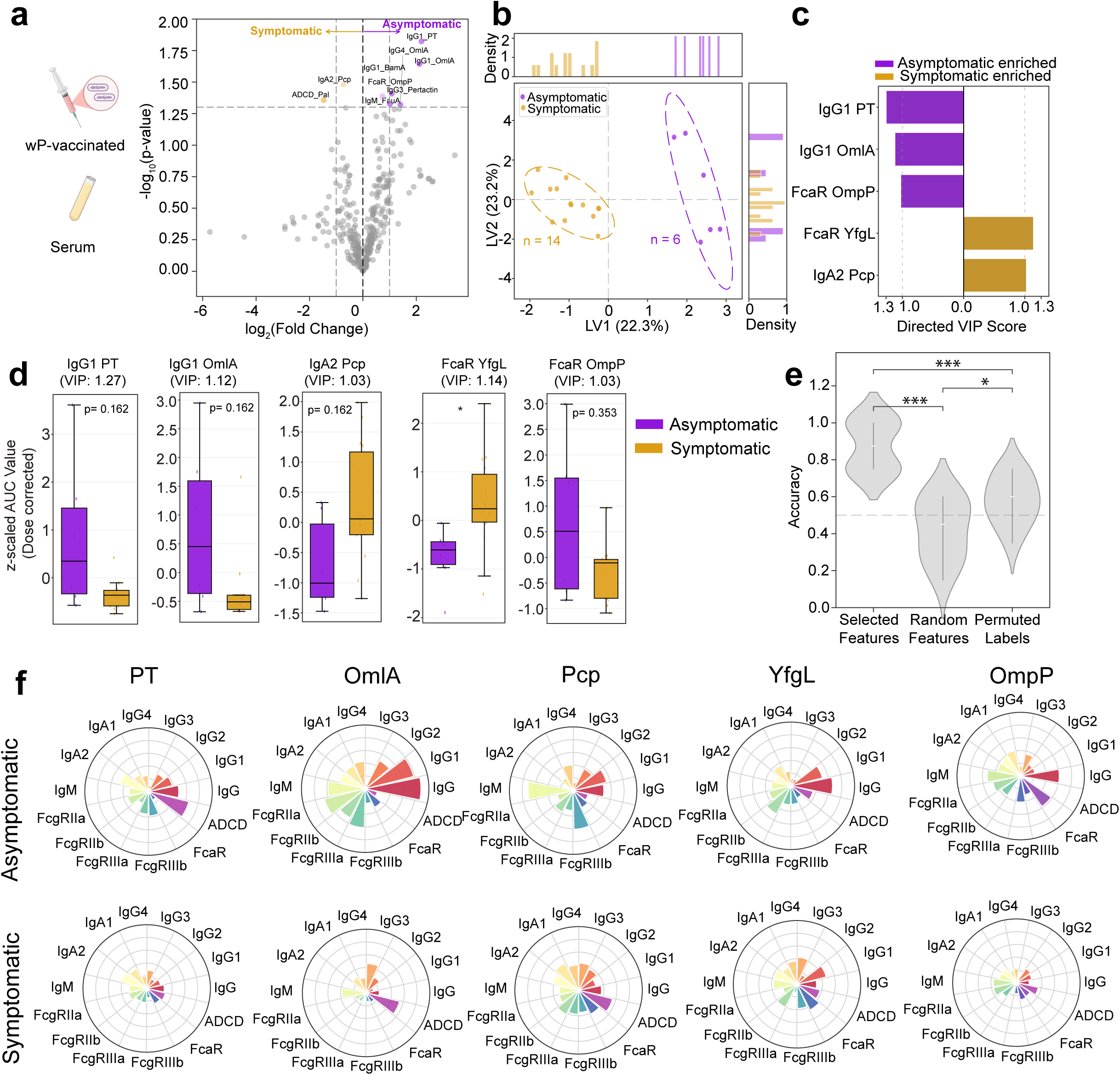
Serum antibody correlates of B. pertussis symptomology in wP-vaccinees. **(a)** Volcano plot showing pairwise comparison depicting serum antibody features between wP vaccinated asymptomatic (purple) and symptomatic (yellow) participants following controlled human infection. Features are plotted as log₂ fold change (dose-corrected AUC day 1 to 7) versus –log₁₀ (p value). The horizontal dashed line indicates p = 0.05, and the vertical dashed line denotes the manually selected significant threshold. **(b)** PLS-DA scores plot demonstrating separation between asymptomatic (purple) or symptomatic (yellow) participants based on LASSO-selected serum antibody profiles (dose-corrected AUC day 1 to 7). Each dot represents an individual participant; the number of individuals (n) is indicated; ellipses denote 75% confidence regions assuming a multivariate t distribution. Marginal density plots for LV1 and LV2 are shown. **(c)** Directed VIP scores highlighting top discriminatory serum features (VIP > 1.0) enriched in asymptomatic (purple) or symptomatic (yellow) individuals. (d) Boxplots representing medians, IQRs, minima, and maxima of representative discriminative features (VIP >1.0, panel c) comparing asymptomatic (purple) or symptomatic (yellow) individuals; the number of individuals is the same as in panel **(b)**. The measurements were dose-corrected and z-scored for AUC day 1 to 7. Significances were assessed using Mann–Whitney U tests and corrected for multiple testing using the Benjamini–Hochberg procedure. Asterisks indicate adjusted P values with *P < 0.05, **P < 0.01, ***P < 0.001. (e) The model performance was validated using permutation tests. Classification accuracy is shown for models built using selected features (panel c), random features, or permuted labels. Violin plots show the distribution from 100 repetitions for each model type. P values represent the median exact permutation-derived P value across repetitions. Significances were assessed using Mann–Whitney U tests and corrected for multiple testing using the Benjamini–Hochberg procedure. Asterisks indicate adjusted P values with *P < 0.05, **P < 0.01, ***P < 0.001. (f) Radar/Polar plots depicting mean percentile ranks of antigen-specific antibody feature distributions for representative discriminative antigens (VIP >1.0, panel c) in non-infected (top) versus infected (bottom) individuals. Percentile rank scores were determined for each antibody feature across all individuals.

**Figure S4.**
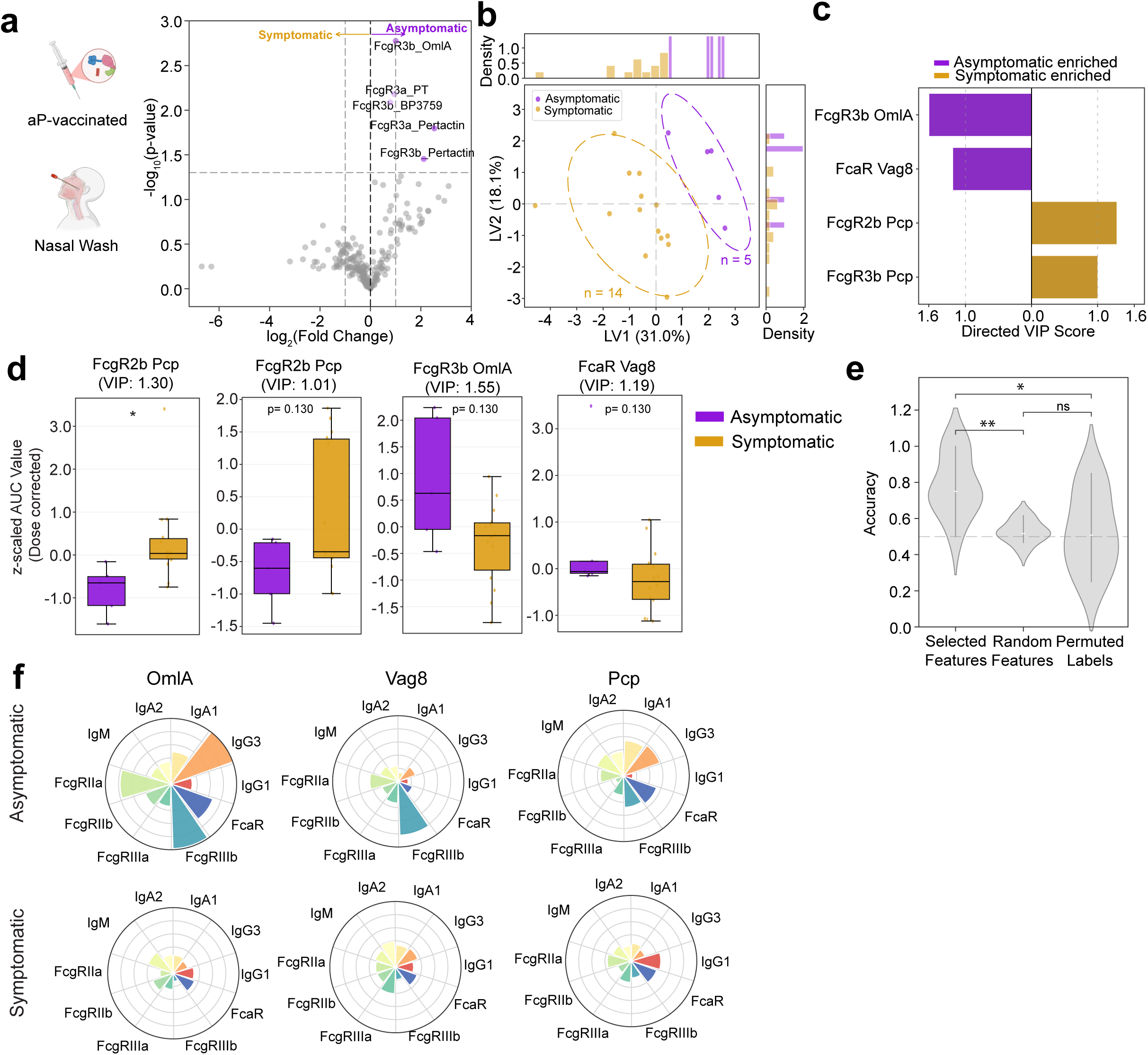
Mucosal antibody correlates of B. pertussis symptomology in aP-vaccinees. **(a)** Volcano plot showing pairwise comparison depicting nasal wash antibody features between aP vaccinated asymptomatic (purple) and symptomatic (yellow) participants following controlled human infection. Features are plotted as log₂ fold change (dose-corrected AUC day 1 to 7) versus –log₁₀ (p value). The horizontal dashed line indicates p = 0.05, and the vertical dashed line denotes the manually selected significant threshold. **(b)** PLS-DA scores plot demonstrating separation between asymptomatic (purple) or symptomatic (yellow) participants based on LASSO-selected nasal wash antibody profiles (dose-corrected AUC day 1 to 7). Each dot represents an individual participant; the number of individuals (n) is indicated; ellipses denote 75% confidence regions assuming a multivariate t distribution. Marginal density plots for LV1 and LV2 are shown. **(c)** VIP scores highlighting top discriminatory nasal wash features (VIP > 1.0) enriched in asymptomatic (purple) or symptomatic (yellow) individuals. (d) Boxplots representing medians, IQRs, minima, and maxima of representative discriminative features (VIP >1.0, panel c) comparing asymptomatic (purple) or symptomatic (yellow) individuals; the number of individuals is the same as in panel **(b)**. The measurements were dose-corrected and z-scored for AUC day 1 to 7. Significances were assessed using Mann–Whitney U tests and corrected for multiple testing using the Benjamini–Hochberg procedure. Asterisks indicate adjusted P values with *P < 0.05, **P < 0.01, ***P < 0.001. (e) The model performance was validated using permutation tests. Classification accuracy is shown for models built using selected features (panel c), random features, or permuted labels. Violin plots show the distribution from 100 repetitions for each model type. P values represent the median exact permutation-derived P value across repetitions. Significances were assessed using Mann–Whitney U tests and corrected for multiple testing using the Benjamini–Hochberg procedure. Asterisks indicate adjusted P values with *P < 0.05, **P < 0.01, ***P < 0.001. (f) Radar/Polar plots depicting mean percentile ranks of antigen-specific antibody feature distributions for representative discriminative antigens (VIP >1.0, panel c) in non-infected (top) versus infected (bottom) individuals. Percentile rank scores were determined for each antibody feature across all individuals.

**Figure S5.**
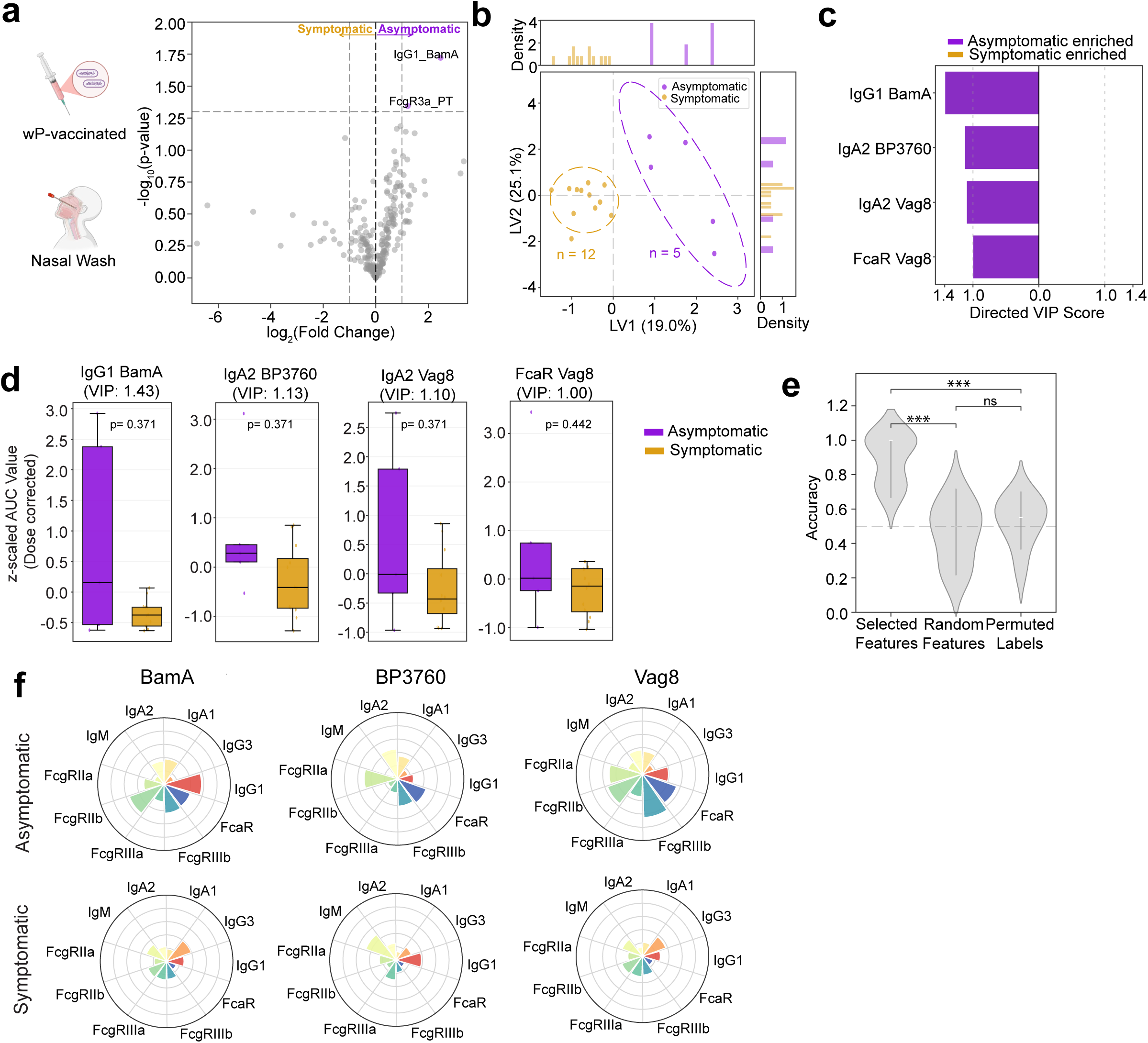
Mucosal antibody correlates of B. pertussis symptomology in wP-vaccinees. **(a)** Volcano plot showing pairwise comparison depicting nasal wash antibody features between wP vaccinated asymptomatic (purple) and symptomatic (yellow) participants following controlled human infection. Features are plotted as log₂ fold change (dose-corrected AUC day 1 to 7) versus –log₁₀ (p value). The horizontal dashed line indicates p = 0.05, and the vertical dashed line denotes the manually selected significant threshold. **(b)** PLS-DA scores plot demonstrating separation between asymptomatic (purple) or symptomatic (yellow) participants based on LASSO-selected nasal wash antibody profiles (dose-corrected AUC day 1 to 7). Each dot represents an individual participan; the number of individuals (n) is indicated t; ellipses denote 75% confidence regions assuming a multivariate t distribution. Marginal density plots for LV1 and LV2 are shown. **(c)** VIP scores highlighting top discriminatory nasal wash features (VIP > 1.0) enriched in asymptomatic (purple) or symptomatic (yellow) individuals. (d) Boxplots representing medians, IQRs, minima, and maxima of representative discriminative features (VIP >1.0, panel c) comparing asymptomatic (purple) or symptomatic (yellow) individuals; the number of individuals is the same as in panel **(b)**. The measurements were dose-corrected and z-scored for AUC day 1 to 7. Significances were assessed using Mann–Whitney U tests and corrected for multiple testing using the Benjamini–Hochberg procedure. Asterisks indicate adjusted P values with *P < 0.05, **P < 0.01, ***P < 0.001. (e) The model performance was validated using permutation tests. Classification accuracy is shown for models built using selected features (panel c), random features, or permuted labels. Violin plots show the distribution from 100 repetitions for each model type. P values represent the median exact permutation-derived P value across repetitions. Significances were assessed using Mann–Whitney U tests and corrected for multiple testing using the Benjamini–Hochberg procedure. Asterisks indicate adjusted P values with *P < 0.05, **P < 0.01, ***P < 0.001. (f) Radar/Polar plots depicting mean percentile ranks of antigen-specific antibody feature distributions for representative discriminative antigens (VIP >1.0, panel c) in non-infected (top) versus infected (bottom) individuals. Percentile rank scores were determined for each antibody feature across all individuals.

**Figure S6.**
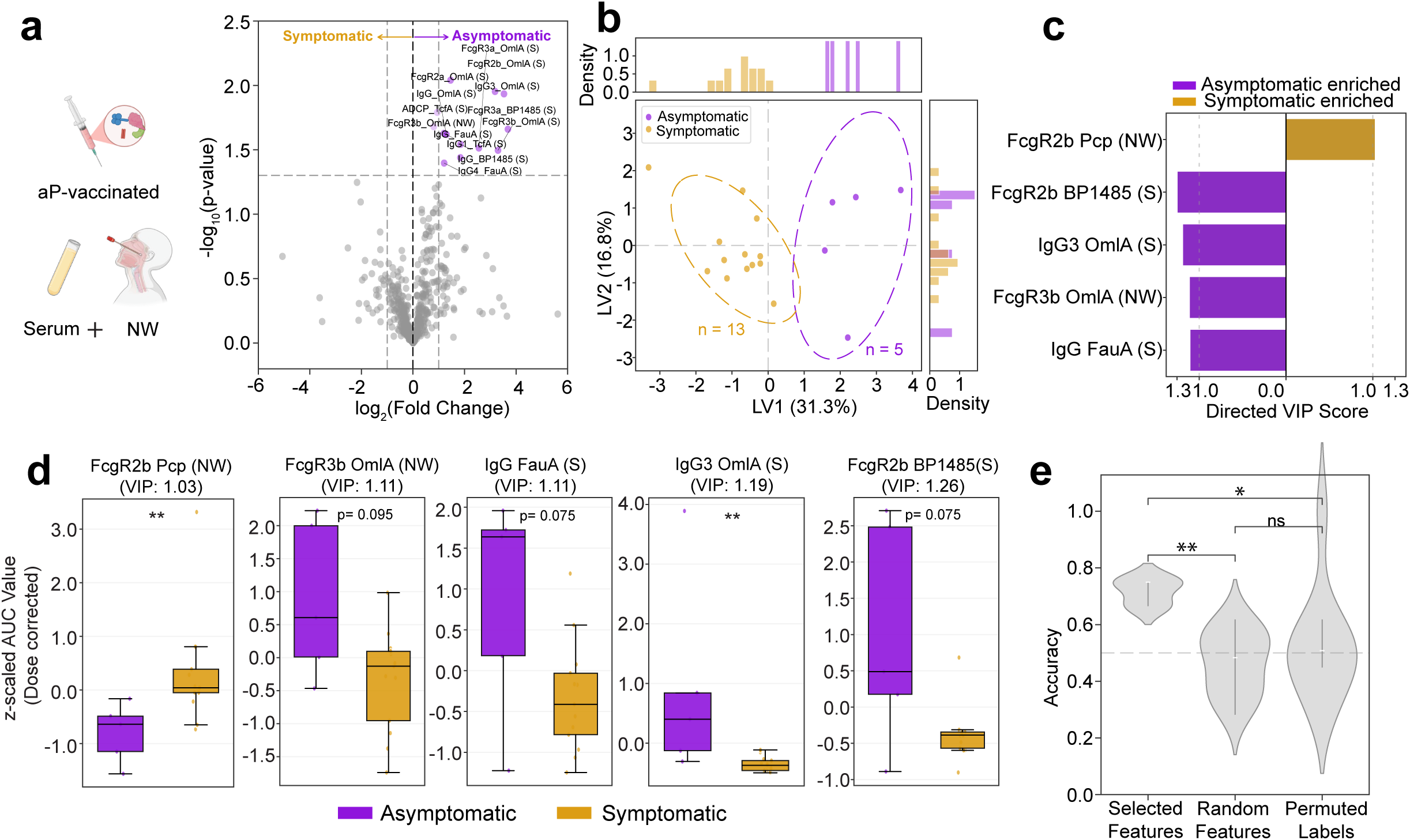
Serum and mucosal antibody correlates of B. pertussis symptomology in aP-vaccinees. **(a)** Volcano plot showing pairwise comparison depicting serum and nasal wash antibody features between aP vaccinated asymptomatic (purple) and symptomatic (yellow) participants following controlled human infection. Features are plotted as log₂ fold change (dose-corrected AUC day 1 to 7) versus –log₁₀ (p value). The horizontal dashed line indicates p = 0.05, and the vertical dashed line denotes the manually selected significant threshold. **(b)** PLS-DA scores plot demonstrating separation between asymptomatic (purple) or symptomatic (yellow) participants based on LASSO-selected serum and nasal wash antibody profiles (dose-corrected AUC day 1 to 7). Each dot represents an individual participant; the number of individuals (n) is indicated; ellipses denote 75% confidence regions assuming a multivariate t distribution. Marginal density plots for LV1 and LV2 are shown. **(c)** VIP scores highlighting top discriminatory antibody features (VIP > 1.0) enriched in asymptomatic (purple) or symptomatic (yellow) individuals. (d) Boxplots representing medians, IQRs, minima, and maxima of representative discriminative features (VIP >1.0, panel c) comparing asymptomatic (purple) or symptomatic (yellow) individuals; the number of individuals is the same as in panel **(b)**. The measurements were dose-corrected and z-scored for AUC day 1 to 7. Significances were assessed using Mann–Whitney U tests and corrected for multiple testing using the Benjamini–Hochberg procedure. Asterisks indicate adjusted P values with *P < 0.05, **P < 0.01, ***P < 0.001. (e) The model performance was validated using permutation tests. Classification accuracy is shown for models built using selected features (panel c), random features, or permuted labels. Violin plots show the distribution from 100 repetitions for each model type. P values represent the median exact permutation-derived P value across repetitions. Significances were assessed using Mann–Whitney U tests and corrected for multiple testing using the Benjamini–Hochberg procedure. Asterisks indicate adjusted P values with *P < 0.05, **P < 0.01, ***P < 0.001.

**Figure S7.**
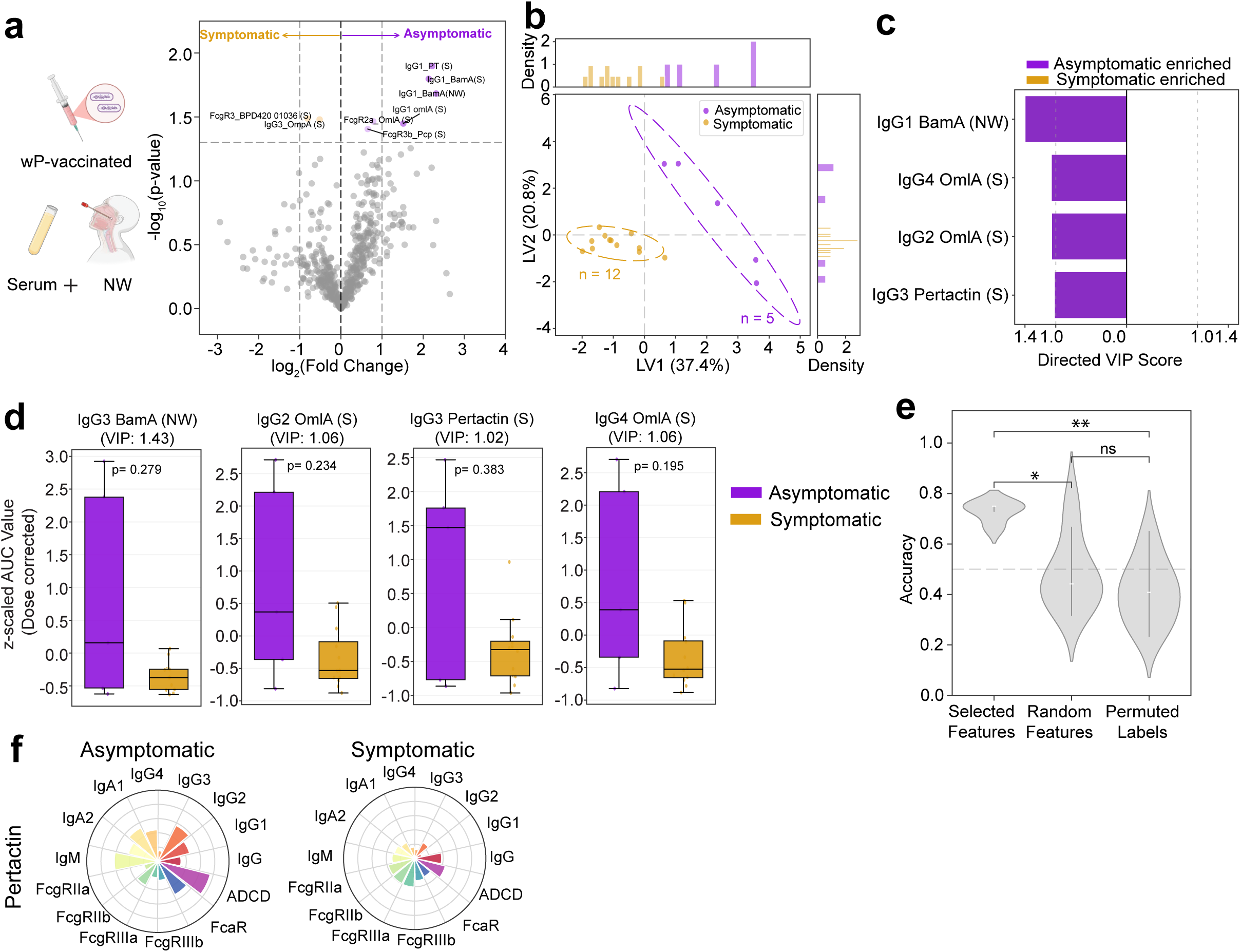
Serum and mucosal antibody correlates of B. pertussis symptomology in wP-vaccinees. **(a)** Volcano plot showing pairwise comparison depicting serum and nasal wash antibody features between wP vaccinated asymptomatic (purple) and symptomatic (yellow) participants following controlled human infection. Features are plotted as log₂ fold change (dose-corrected AUC day 1 to 7) versus –log₁₀ (p value). The horizontal dashed line indicates p = 0.05, and the vertical dashed line denotes the manually selected significant threshold. **(b)** PLS-DA scores plot demonstrating separation between asymptomatic (purple) or symptomatic (yellow) participants based on LASSO-selected serum and nasal wash antibody profiles (dose-corrected AUC day 1 to 7). Each dot represents an individual participant; the number of individuals (n) is indicated; ellipses denote 75% confidence regions assuming a multivariate t distribution. Marginal density plots for LV1 and LV2 are shown. **(c)** VIP scores highlighting top discriminatory antibody features (VIP > 1.0) enriched in asymptomatic (purple) or symptomatic (yellow) individuals. (d) Boxplots representing medians, IQRs, minima, and maxima of representative discriminative features (VIP >1.0, panel c) comparing asymptomatic (purple) or symptomatic (yellow) individuals; the number of individuals is the same as in panel **(b)**. The measurements were dose-corrected and z-scored for AUC day 1 to 7. Significances were assessed using Mann–Whitney U tests and corrected for multiple testing using the Benjamini–Hochberg procedure. Asterisks indicate adjusted P values with *P < 0.05, **P < 0.01, ***P < 0.001. (e) The model performance was validated using permutation tests. Classification accuracy is shown for models built using selected features (panel c), random features, or permuted labels. Violin plots show the distribution from 100 repetitions for each model type. P values represent the median exact permutation-derived P value across repetitions. Significances were assessed using Mann–Whitney U tests and corrected for multiple testing using the Benjamini–Hochberg procedure. Asterisks indicate adjusted P values with *P < 0.05, **P < 0.01, ***P < 0.001. (f) Radar/Polar plots depicting mean percentile ranks of antigen-specific antibody feature distributions for representative discriminative antigens (VIP >1.0, panel c) in non-infected (top) versus infected (bottom) individuals. Percentile rank scores were determined for each antibody feature across all individuals.

**Figure S8.**
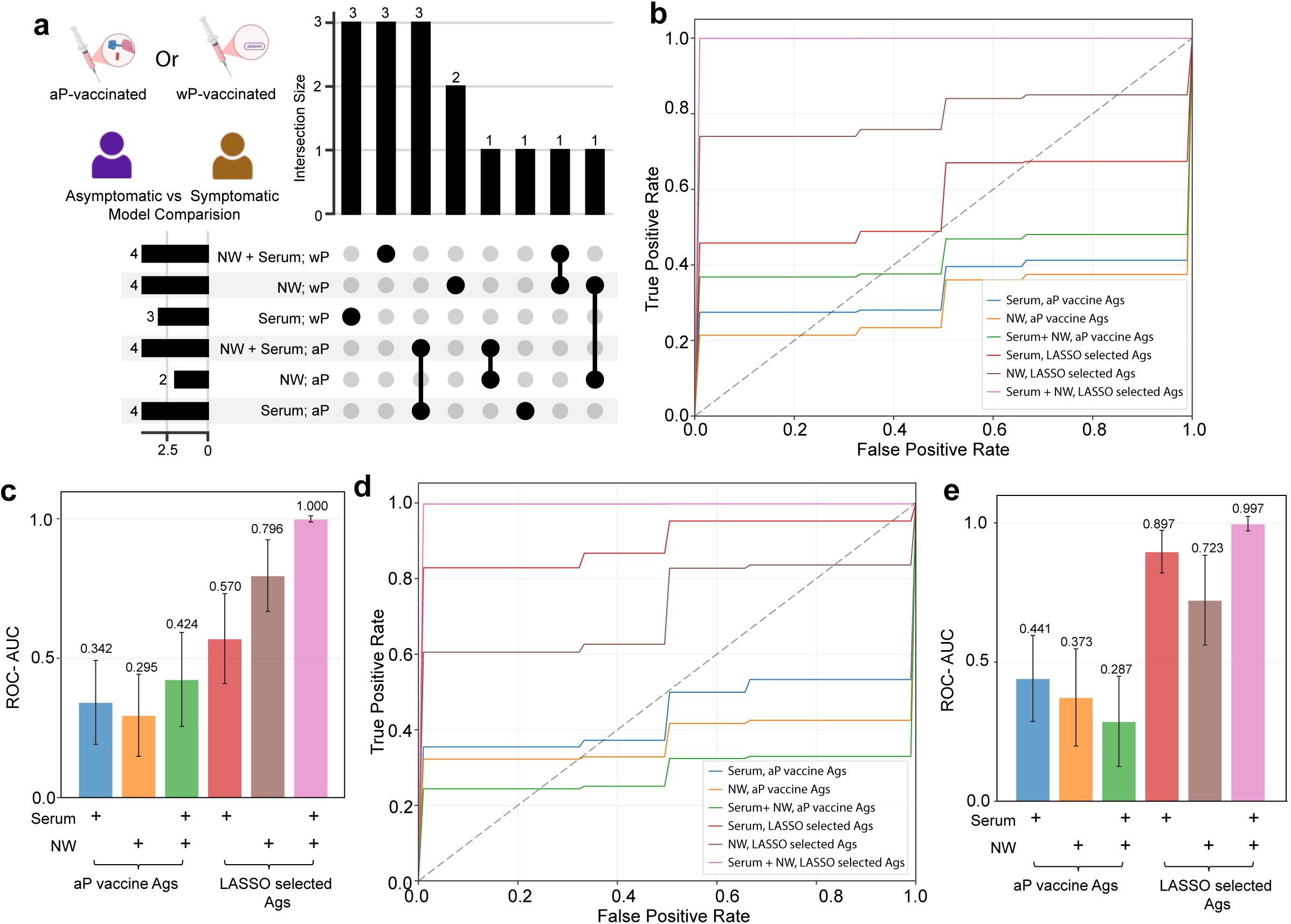
Integrated serum and mucosal antibody features improve discrimination of symptomatic B. pertussis cases in wP- and aP- vaccinees. (a) UpSet plot showing the overlap of discriminative antigen sets derived from LASSO/PLSDA models using antibody features in serum, nasal wash, or combined serum + nasal wash in wP- and aP- vaccinated participants. (b,d) Receiver operating characteristic (ROC) curves comparing model performance for classifying asymptomatic versus symptomatic participants using serum alone, nasal wash alone, or combined serum + nasal wash antibody features across antigen classes (aP vaccine antigens, LASSO-selected antigens, and microbiome antigens) in aP-vaccinated (b) and wP-vaccinated (e) participants. (c,e) Bar plot summarizing ROC–AUC values for each model in aP-vaccinated (c) and wP-vaccinated (e) participants. Bars represent mean ROC–AUC ± S.D. (d,g).

## Supplemental Table

**Table S1. Metadata and antigen panel of systems serology.** PBS subtracted values for distinct antigen-specific antibody features.

**Table S2. Features enriched in various categories as identified by univariate analysis (volcano plots)**

**Table S3. Serum correlates of protection from infection and symptomatology identified by LASSO-PLSDA**

**Table S4. Nasal wash correlates of protection from infection and symptomatology identified by LASSO-PLSDA**

**Table S5. Combined serum and nasal wash correlates of protection from infection and symptomatology identified by LASSO-PLSDA**

**Table S6. Non-infected VIP features identified across sample types in wP-vaccinated participants**

**Table S7. Asymptomatic VIP features identified across sample types in aP- and wP-vaccinated participants**

## Materials and Methods

### Study Cohort and Sample Information

Serum and nasal wash samples were obtained from healthy adult volunteers (18-40 years) enrolled in a controlled human infection model (CHIM) of *Bordetella pertussis* conducted at the Canadian Center for Vaccinology (CCfV), Dalhousie University. This open-label, phase 1, dose-escalation clinical trial was designed to establish a safe and reproducible human challenge model using the pertactin-producing *B. pertussis* D420 isolate administered intranasally, published elsewhere^14^. In brief, eligible participants were screened to exclude individuals with underlying chronic medical conditions, immunodeficiency, pregnancy, abnormal laboratory findings, recent respiratory illness, or significant psychiatric disease. Individuals with recent pertussis vaccination or serologic evidence of high baseline pertussis immunity (anti-PT titers >20 EU/ml) were excluded^14^. Participants were challenged with a single intranasal inoculation of *B. pertussis* D420 across escalating dose cohorts (10^3^-10^8^ CFU) to determine the human infectious dose associated with bacterial colonization and mild symptomatic infection.

Following challenge, participants were monitored longitudinally during an inpatient phase and subsequent outpatient follow-up. Serum and nasal wash samples were collected at baseline prior to challenge and at multiple post-challenge time points extending through day 180. Nasopharyngeal colonization was assessed by serial culture and quantitative PCR, with bacterial detection evaluated beginning on day 6 post-challenge to account for incubation and avoid confounding by residual inoculum. Infection status was defined by detection of *B. pertussis* by culture or PCR, and symptom status was determined based on prospectively collected solicited clinical symptoms. Participants were classified into outcome groups including non-infected (no bacterial detection), infected without symptoms i.e. asymptomatic infection (bacterial detection but no solicited symptoms), and infected with symptoms i.e. symptomatic infection (bacterial detection with symptoms) using a blinded adjudication process and an automated algorithm integrating bacterial shedding and symptom criteria. Bacterial detection was defined by ≥1 positive culture or <u>>3</u> positive PCR from day 6 post challenge to first day of treatment. Symptomatic cases had confirmed infection status meeting the following complement: 2 or more symptoms, at least one of which must be a respiratory specific symptom.

All participants received azithromycin eradication therapy per protocol, regardless of clinical outcome. All participants provided written informed consent under protocols approved by Health Canada, the U.S. Food and Drug Administration, and local institutional review boards. 4 Cohorts challenged with 10^6^ to 10^8^ CFU were deidentified prior to shipment to the Ragon Institute of Massachusetts General Brigham, MIT and Harvard for systems serology analysis. Investigators performing immunologic assays remained blinded until completion of data acquisition, quality control, and integration.

### Luminex-Based Systems Serology

Custom Luminex MagPlex bead–based multiplex assays were used to quantify antigen-specific antibody isotypes and subclasses (IgG, IgG1–4, IgA1, IgA2, and IgM) and Fc receptor binding (FcγR2a, FcγR2b, FcγR3a, FcγR3b, FcαR) across a curated panel of control, microbiome-derived, pertussis, and TDAP antigens (Table S1). Carboxylated Luminex microspheres were covalently coupled to protein antigens using two-step EDC/sulfo-NHS chemistry or to polysaccharides using DMTMM activation, followed by blocking and storage in PBS. Serum samples were heat-inactivated at 56 °C for 30 min, and assay conditions were optimized over dilution curves to ensure measurements within the linear dynamic range. 5ul of diluted serum samples (1:200 for IgG; 1:50 for IgG1 and IgG3; 1:10 for IgG2, IgG4, IgA1, IgA2 and FcαR; 1:5 for IgM; 1:40 for FcγRs) and nasal wash samples (10ul for IgG, IgG1, IgG3 and IgM; 15ul for IgG2 and IgG4; 5ul for IgA1, IgA2, FcγRs and FcαR) were incubated with antigen-coupled beads overnight at 4°C.

Antibody isotype and subclass detection was performed using PE-conjugated secondary antibodies (SouthernBiotech). Fc receptor binding was assessed using biotinylated recombinant human Fcγ receptors produced at the Duke University Protein Production Core using BirA-mediated biotinylation, followed by detection with streptavidin-PE (PhycoLink). All secondary incubations were performed for 1 h at room temperature.

Bead-associated fluorescence was acquired on a xMAP Intelliflex system (Luminex Corp.). All samples were assayed in duplicate, values were averaged, and background signal from PBS controls was subtracted prior to downstream analysis.

### Quantification and statistical analysis

All assays were run in duplicated and measurements were averaged for each sample and PBS subtracted. Group comparisons were performed using Mann-Whitney U tests. Dose correction were conducted using linear regression with dose as a covariate. Volcano plots shows log_2_ fold change (log_2_FC) vs. statistical significance (−log₁₀p-value), with p < 0.05 and log_2_FC as the nominal threshold. Correlation analyses used Spearman’s R. All analyses were conducted in Python (Version 3.13.1).

### Antibody-Dependent Cellular Phagocytosis (ADCP) and Antibody-Dependent Neutrophil Phagocytosis (ADNP)

Bead-based assays were used to quantify antibody-dependent cellular phagocytosis (ADCP) and antibody-dependent neutrophil phagocytosis (ADNP), as previously described^31^. Antigens, selected based on univariate analysis of non-infected versus infected and asymptomatic versus symptomatic, were biotinylated using Sulfo-NHS-LC-LC biotin (Thermo Fisher Scientific, Waltham, MA, USA) according to the manufacturer’s instructions. For multiplexing, antigens were coupled to custom-made fluorescent NeutrAvidin beads (Thermo Fisher Scientific). Bead mixtures were incubated with diluted serum samples for 2 h at 37 °C to allow immune complex formation. For ADCP, THP-1 monocytes (ATCC) were added to immune complexes at a concentration of 1.25 × 10⁵ cells/mL and incubated for 16 h at 37 °C. For ADNP, primary human neutrophils were isolated from whole blood by ACK red blood cell lysis (Quality Biological). Isolated neutrophils (50,000 cells per well) were incubated with immune complexes for 2 h at 37 °C. Following incubation, neutrophils were stained with anti-CD66b Pacific Blue antibody (BioLegend) and fixed with 4% paraformaldehyde (Santa Cruz Biotechnology). Samples were acquired by flow cytometry using an iQue Screener PLUS (Intellicyt) equipped with an S-Lab 384-well plate handling robot (PAA).

For ADCP analysis, events were gated on singlets followed by bead-positive cells. For ADNP, neutrophils were identified as CD66b⁺ events and further gated on bead-positive populations. A phagocytosis score was calculated for both ADCP and ADNP as: (percentage of bead-positive cells) x (MFI of bead-positive cells) divided by 10,000. Each sample was assayed in two independent technical replicates, and replicate values were averaged after subtraction of background signal from PBS controls prior to downstream analysis.

### Antibody-Dependent Complement Deposition (ADCD)

Antibody-dependent complement deposition (ADCD) was assessed, as previously described^32^. Briefly, antigen-coupled Luminex beads were incubated with heat-inactivated serum samples dilute for 1 h at 37 °C to allow immune complex formation. Lyophilized guinea pig complement (Cedarlane) was reconstituted according to the manufacturer’s instructions and diluted in gelatin veronal buffer containing calcium and magnesium (GVB++). Complement was added to immune complexes and incubated for 20–30 min at 37 °C to permit C3 deposition. Following incubation, deposited C3 was detected using a PE- or fluorescein-conjugated anti–guinea pig C3 detection antibody (MP Biomedicals). Beads were washed and acquired on a Luminex xMAP Intelliflex system. Complement deposition was quantified as the median fluorescence intensity (MFI) of C3 signal for each antigen-coupled bead region. MFI from technical duplicates were averaged after subtraction of background signal from PBS controls.

### Feature Selection via Stability-Based LASSO

Prior to multivariate analysis, all antibody antibody responses were calculated as AUC values across days −1, 1, and 3 (or 1, 3, and 7), dose-corrected using linear regression, log-transformed (where necessary) and Z-score normalized using the *StandardScaler* function from scikit-learn. To reduce overfitting and identify robust immune correlates, a stability-based Least Absolute Shrinkage and Selection Operator (LASSO) framework was implemented.

A five-fold cross-validation strategy was used, in which the dataset was partitioned into five equally sized folds. For each iteration, four folds (80% of samples) were used to train the LASSO model, while the remaining fold (20%) was used as an independent test set. Model performance was evaluated using classification accuracy on the held-out fold. This process was repeated such that each fold served as the test set once. Feature stability was assessed across repeated iterations, and only features selected in ≥90% of runs were retained for downstream modeling.

### Classification and Visualization Using PLS-DA

Features selected by the stability LASSO were used to construct a Partial Least Squares Discriminant Analysis (PLS-DA) model using the *PLSRegression* module in scikit-learn, with selected antibody features regressed against binary outcome labels. Model performance was assessed using five-fold stratified cross-validation, and receiver operating characteristic (ROC) curves were generated using *sklearn.metrics.roc_curve*.

To evaluate model specificity, performance of the true PLS-DA model was compared against two negative control models: (1) a model built using an equal number of randomly selected features, and (2) a model built using the selected features with permuted class labels. Statistical significance between model accuracies was assessed using the Mann–Whitney U test (*scipy.stats.mannwhitneyu*).

The contribution of individual features to class separation was quantified using Variable Importance in Projection (VIP) scores, with features exhibiting VIP > 1.0 considered highly influential. Directionality of feature enrichment was visualized using directional VIP bar plots. Sample-wise scores from the first two latent variables (LV1 and LV2) were plotted to visualize group separation, with confidence ellipses representing group-level dispersion.

### Receiver Operating Characteristic (ROC) Analysis for Model Performance

Receiver operating characteristic (ROC) analysis was performed to evaluate the ability of models to discriminate groups. Separate classification models were constructed using antibody features derived from serum alone, nasal wash (NW) alone, or combined serum and nasal wash measurements. Feature sets were grouped into three predefined antigen classes: acellular pertussis (aP) vaccine antigens, LASSO-selected antigens, and microbiome-associated antigens.

For each feature set and compartment (serum, NW, or serum + NW), Partial Least Squares Discriminant Analysis (PLS-DA) models were trained using five-fold stratified cross-validation. At each iteration, predicted class probabilities from the held-out test fold were used to generate ROC curves, plotting true positive rate against false positive rate across classification thresholds. ROC curves were generated using the *roc_curve* function from *sklearn.metrics*, and the area under the ROC curve (AUC) was calculated to quantify model performance.

ROC curves represent the aggregated performance across cross-validation folds for each model configuration. A diagonal reference line indicates performance equivalent to random classification. Comparative ROC analysis (Mann-Whitney U tests with Benjamini-Hochberg false discovery rate (FDR) correction) was used to assess the relative discriminatory power of serum, nasal wash, and combined antibody features across antigen classes. These analyses demonstrate improved classification performance when LASSO-selected features and combined serum + nasal wash data are incorporated into the model.

## Data Availability

All data produced in the present study are available upon reasonable request to the authors

## Acknowledgments

We thank T. Ragon and S. Ragon and the Ragon Institute of MGH, MIT and Harvard for support. We thank Yihui Wang and Tod Merkel for critical discussions.

## Author contributions

M.Z.K, G.A., and B.J conceptualized the study. Luminex-based measurements of isotype and subclass concentrations, FcR binding, avidity, and all functional assays were performed by M.Z.K, Y.K, T.S. and A.E.L. E.K. performed all analysis and created all data visualizations under the supervision of B.J, G.A. and D.L. Samples were collected and metadata was curated by S.H. Project administration was performed by L.F, M.C, S.H, M.E. The manuscript was drafted by M.Z.K, Y.K, E.K, G.A, and B.J and reviewed and edited by all authors.

## Data and materials availability

Metadata associated with this study is provided in Table S1.

## References

1. Fullen AR, Yount KS, Dubey P, Deora R. Whoop! There it is: The surprising resurgence of pertussis. PLoS Pathog 2020; 16(7): e1008625.

2. Burdin N, Handy LK, Plotkin SA. What Is Wrong with Pertussis Vaccine Immunity? The Problem of Waning Effectiveness of Pertussis Vaccines. Cold Spring Harb Perspect Biol 2017; 9(12).

3. Warfel JM, Zimmerman LI, Merkel TJ. Acellular pertussis vaccines protect against disease but fail to prevent infection and transmission in a nonhuman primate model. Proc Natl Acad Sci U S A 2014; 111(2): 787–92.

4. Kapil P, Merkel TJ. Pertussis vaccines and protective immunity. Curr Opin Immunol 2019; 59: 72–8.

5. Barbic J, Leef MF, Burns DL, Shahin RD. Role of gamma interferon in natural clearance of Bordetella pertussis infection. Infect Immun 1997; 65(12): 4904–8.

6. Leef M, Elkins KL, Barbic J, Shahin RD. Protective immunity to Bordetella pertussis requires both B cells and CD4(+) T cells for key functions other than specific antibody production. J Exp Med 2000; 191(11): 1841–52.

7. Allen AC, Wilk MM, Misiak A, Borkner L, Murphy D, Mills KHG. Sustained protective immunity against Bordetella pertussis nasal colonization by intranasal immunization with a vaccine-adjuvant combination that induces IL-17-secreting T(RM) cells. Mucosal Immunol 2018; 11(6): 1763–76.

8. Ross PJ, Sutton CE, Higgins S, et al. Relative contribution of Th1 and Th17 cells in adaptive immunity to Bordetella pertussis: towards the rational design of an improved acellular pertussis vaccine. PLoS Pathog 2013; 9(4): e1003264.

9. van der Lee S, Sanders EAM, Berbers GAM, Buisman AM. Whole-cell or acellular pertussis vaccination in infancy determines IgG subclass profiles to DTaP booster vaccination. Vaccine 2018; 36(2): 220–6.

10. Warfel JM, Merkel TJ. Bordetella pertussis infection induces a mucosal IL-17 response and long-lived Th17 and Th1 immune memory cells in nonhuman primates. Mucosal Immunol 2013; 6(4): 787–96.

11. Wilk MM, Misiak A, McManus RM, Allen AC, Lynch MA, Mills KHG. Lung CD4 Tissue-Resident Memory T Cells Mediate Adaptive Immunity Induced by Previous Infection of Mice with Bordetella pertussis. J Immunol 2017; 199(1): 233–43.

12. Gregg KA, Wang Y, Warfel J, et al. Antigen Discovery for Next-Generation Pertussis Vaccines Using Immunoproteomics and Transposon-Directed Insertion Sequencing. J Infect Dis 2023; 227(4): 583–91.

13. Chung AW, Kumar MP, Arnold KB, et al. Dissecting Polyclonal Vaccine-Induced Humoral Immunity against HIV Using Systems Serology. Cell 2015; 163(4): 988–98.

14. ElSherif MS, Redden KL, Langley JM, et al. A controlled human infection model for symptomatic pertussis in North America using the pertactin-producing clinical isolate D420. MedRxiv 2026.

15. Mettelman RC, Allen EK, Thomas PG. Mucosal immune responses to infection and vaccination in the respiratory tract. Immunity 2022; 55(5): 749–80.

16. von Messling VA, Griffin DE. How respiratory viruses overcome mucosal defenses and exploit the unique environment of the respiratory tract. Curr Opin Virol 2012; 2(3): 221–4.

17. Zhao M, Zhou L, Wang S. Immune crosstalk between respiratory and intestinal mucosal tissues in respiratory infections. Mucosal Immunol 2025; 18(3): 501–8.

18. Marcellini V, Piano Mortari E, Fedele G, et al. Protection against Pertussis in Humans Correlates to Elevated Serum Antibodies and Memory B Cells. Front Immunol 2017; 8: 1158.

19. Aase A, Herstad TK, Merino S, et al. Opsonophagocytic activity and other serological indications of Bordetella pertussis infection in military recruits in Norway. Clin Vaccine Immunol 2007; 14(7): 855–62.

20. Goldsmith JA, Nguyen AW, Wilen RE, Wijagkanalan W, McLellan JS, Maynard JA. Structural Basis for Antibody Neutralization of Pertussis Toxin. bioRxiv 2024.

21. Granstrom M, Granstrom G, Gillenius P, Askelof P. Neutralizing antibodies to pertussis toxin in whooping cough. J Infect Dis 1985; 151(4): 646–9.

22. Sutherland JN, Chang C, Yoder SM, Rock MT, Maynard JA. Antibodies recognizing protective pertussis toxin epitopes are preferentially elicited by natural infection versus acellular immunization. Clin Vaccine Immunol 2011; 18(6): 954–62.

23. Weingart CL, Mobberley-Schuman PS, Hewlett EL, Gray MC, Weiss AA. Neutralizing antibodies to adenylate cyclase toxin promote phagocytosis of Bordetella pertussis by human neutrophils. Infect Immun 2000; 68(12): 7152–5.

24. Zackrisson G, Lagergard T, Trollfors B, Krantz I. Immunoglobulin A antibodies to pertussis toxin and filamentous hemagglutinin in saliva from patients with pertussis. J Clin Microbiol 1990; 28(7): 1502–5.

25. Klein NP, Bartlett J, Fireman B, Rowhani-Rahbar A, Baxter R. Comparative effectiveness of acellular versus whole-cell pertussis vaccines in teenagers. Pediatrics 2013; 131(6): e1716–22.

26. Storsaeter J, Hallander HO, Gustafsson L, Olin P. Levels of anti-pertussis antibodies related to protection after household exposure to Bordetella pertussis. Vaccine 1998; 16(20): 1907–16.

27. Burnham-Marusich AR, Olsen RK, Scarbrough J, et al. Tracheal colonization factor A (TcfA) is a biomarker for rapid and specific detection of Bordetella pertussis. Sci Rep 2020; 10(1): 15002.

28. de Gouw D, de Jonge MI, Hermans PW, et al. Proteomics-identified Bvg-activated autotransporters protect against bordetella pertussis in a mouse model. PLoS One 2014; 9(8): e105011.

29. Oliver DC, Fernandez RC. Antibodies to BrkA augment killing of Bordetella pertussis. Vaccine 2001; 20(1-2): 235–41.

30. Brickman TJ, Armstrong SK. Essential role of the iron-regulated outer membrane receptor FauA in alcaligin siderophore-mediated iron uptake in Bordetella species. J Bacteriol 1999; 181(19): 5958–66.

31. Butler AL, Fallon JK, Alter G. A Sample-Sparing Multiplexed ADCP Assay. Front Immunol 2019; 10: 1851.

32. Kaplonek P, Fischinger S, Cizmeci D, et al. mRNA-1273 vaccine-induced antibodies maintain Fc effector functions across SARS-CoV-2 variants of concern. Immunity 2022; 55(2): 355–65 e4.

